# Automatic assessment of mammographic density using a deep transfer learning method

**DOI:** 10.1101/2022.08.31.22279460

**Authors:** Steven Squires, Elaine Harkness, D. Gareth Evans, Susan M Astley

## Abstract

**Purpose:** Mammographic breast density is one of the strongest risk factors for cancer. Density assessed by radiologists using visual analogue scales has been shown to provide better risk predictions than other methods. Our purpose is to build automated models using deep learning and train on radiologist scores to make accurate and consistent predictions.

**Approach:** We used a data-set of almost 160,000 mammograms each with two independent density scores made by expert medical practitioners. We used two pre-trained deep networks and adapted them to produce feature vectors which were then used for both linear and non-linear regression to make density predictions. We also simulated an “optimal method” which allowed us to compare the quality of our results with a simulated upper bound on performance.

**Results:** Our deep learning method produced estimates with a Root Mean Squared Error (RMSE) of 8.79 ± 0.21. The model estimates of cancer risk perform at a similar level to human experts, within uncertainty bounds. We made comparisons between different model variants and demonstrated the high level of consistency of the model predictions. Our modelled “optimal method” produced image predictions with a RMSE of between 7.98 and 8.90 for cranial caudal images.

**Conclusion:** We demonstrated a new deep learning framework based upon a transfer learning approach to make density estimates based on radiologists’ visual scores. Our approach requires modest computational resources and has the potential to be trained with limited quantities of data.

## 1 Introduction

Studies have shown a strong relationship between breast density and the risk of developing breast cancer.^1–3^ Breast density is generally defined as the proportion of fibro-glandular tissue within the breast, however there are various different methods to estimate this measure and these show different levels of correlation with cancer risk, with breast density assessed subjectively by radiologists demonstrating a stronger link to breast cancer than other methods.^4, 5^ Exactly why this subjective type of measure produces improved performance is not clear, although presumably radiologists utilise years of knowledge and experience to go beyond simple estimates of ratios of dense to non-dense tissue. Their knowledge and experience can be harnessed by training an automated method to produce similar density assessments. If we can accurately measure breast density over time we can also measure whether risk-reducing interventions are working effectively.^6^

Deep learning is now the dominant method used in general image analysis tasks due to the higher accuracy it tends to achieve over more traditional methods.^7^ The main alternative to deep learning is to hand-craft features and then apply traditional machine learning methods. The advantage of deep learning is that the features are automatically extracted from the data itself. This is appealing for breast density estimation as we do not completely understand why subjective expert judgement seems to outperform other methods. Deep learning does have significant downsides, such as the requirement of significant amounts of data and computing power. In addition, the interpretation of results is challenging.^8^

Medical imaging problems often have significantly smaller amounts of data than datasets that deep learning is usually trained on. One of the most commonly used non-medical imaging data-sets is ImageNet^9^ which consists of over a million images and a thousand classes. Conversely, medical imaging data-sets tend to be considered large if they contain tens of thousands of images, with many data-sets consisting of far fewer.^10^ Even with these challenges deep learning is increasingly being used in medical imaging studies^11^ with good outcomes.

There have been many approaches for making breast density estimates. One example^12^ used convolutional neural networks (CNNs) with support vector machines (SVMs) to classify breasts into four different density categories. Other work used unsupervised CNNs at different scales to extract features before finetuning on known labels.^13^ Another method used fuzzed c-means before applying an SVM.^14^ One recent approach^15^ built a deep learning model and trained it from scratch to make estimates of breast density using domain expert labelled images as targets. The authors showed that deep learning models could make estimates that correlate well with the expert labels.

Deep learning can either be performed by designing and training a model from scratch or with a transfer learning approach.^16^ At the moment there is little agreement across the medical imaging field about which option produces better outcomes.^10^ In a field as new as deep learning, with little solid underlying theory and estimates being made on highly complex data-sets, and using complex models, it is difficult to draw strong conclusions about which method to follow. A sensible approach is to test multiple methods on different problems and attempt to determine which models work better in certain situations. A transfer learning method to estimate breast density for low dose mammograms recently showed good performance^17^ albeit on a small data-set; in this paper we will demonstrate that transfer learning using deep networks produces good performance using a large full dose data-set.

We present a transfer learning method based upon two independent deep learning models trained on ImageNet^9^ with regression models trained using a data-set with visual labels produced by domain experts.^18^ These models are combined using a multi-layer-perceptron (MLP) to make a final ensemble prediction. We compare these results with those of a previous method trained on this data-set.^15^ Each image in the data-set was previously assessed by two independent readers which gives us the opportunity to analyse the quality of the labels themselves.^18^ We will show there are challenges with using data with this level of noise both in terms of training but also in how we assess model performance.

The key contributions of this work are:

- We design and implement a transfer-learning based pipeline to make estimates of breast density. Our framework consists of pre-processing, feature extraction, density mapping and final ensemble estimation steps. The overall method produces automated breast density estimates with relatively low computational requirements.
- We analyse the data-set itself to assess how well our model performs. We demonstrate the that the variability of the reader assessment means we are limited in our ability to effectively discriminate between the quality of models once they have reached a certain accuracy. Our framework produces estimates that fall close to this range.

## 2 Data

The data-set is formed of mammogram images with associated density assessments by domain experts (radiologists, advanced practitioner radiographers and breast physicians). The images are from the Predicting Risk Of Cancer At Screening (PROCAS)^18^ study. All images were produced using GE mammography machines and have three different image sizes: 2, 294 × 1, 914; 3, 062 × 2, 394; and 5, 625 × 4, 095. There are four views for each woman: cranial caudal (CC) and mediolateral oblique (MLO) for the left (L) and right (R) breast giving RCC, RMLO, LCC and LMLO views.

In the PROCAS study every image was independently viewed by two domain experts, out of a total pool of 19, who assigned a density value on a visual analogue scale (VAS) between 0 and 100 for each image. Therefore each woman received eight estimates of breast density across both CC and MLO and right and left breasts.

Before performing any pre-processing we removed any images that do not have labels assigned from two readers. In Table 1 we show the number of images at the three different sizes in our dataset. The images are drawn from 39,357 women, although not all the women have the full set of image views. When considering predictions per image we will use the entire data-set but when analysing density estimates per woman we will only consider those women who have all four views. In Figure 1 we show image examples of CC (top row) and MLO (bottom row) images with low, medium and high densities (from left to right) as defined by the average of the two reader scores. These are images after pre-processing (see Algorithm 1 in Section 3.1 for details).

**Table 1.** Number of images at each size and aspect ratio

| Name | Height by width | Number of pixels | Aspect Ratio | Number of Images |
| --- | --- | --- | --- | --- |
| **Small** | $2,294 \times 1,914$ | 4,390,716 | 1 : 1.20 | 57,733 |
| **Medium** | $3,062 \times 2,394$ | 7,330,428 | 1 : 1.28 | 95,184 |
| **Large** | $5,625 \times 4,095$ | 23,034,375 | 1 : 1.37 | 4,055 |
| **All** | - | - | - | 156,972 |

**Fig 1.**
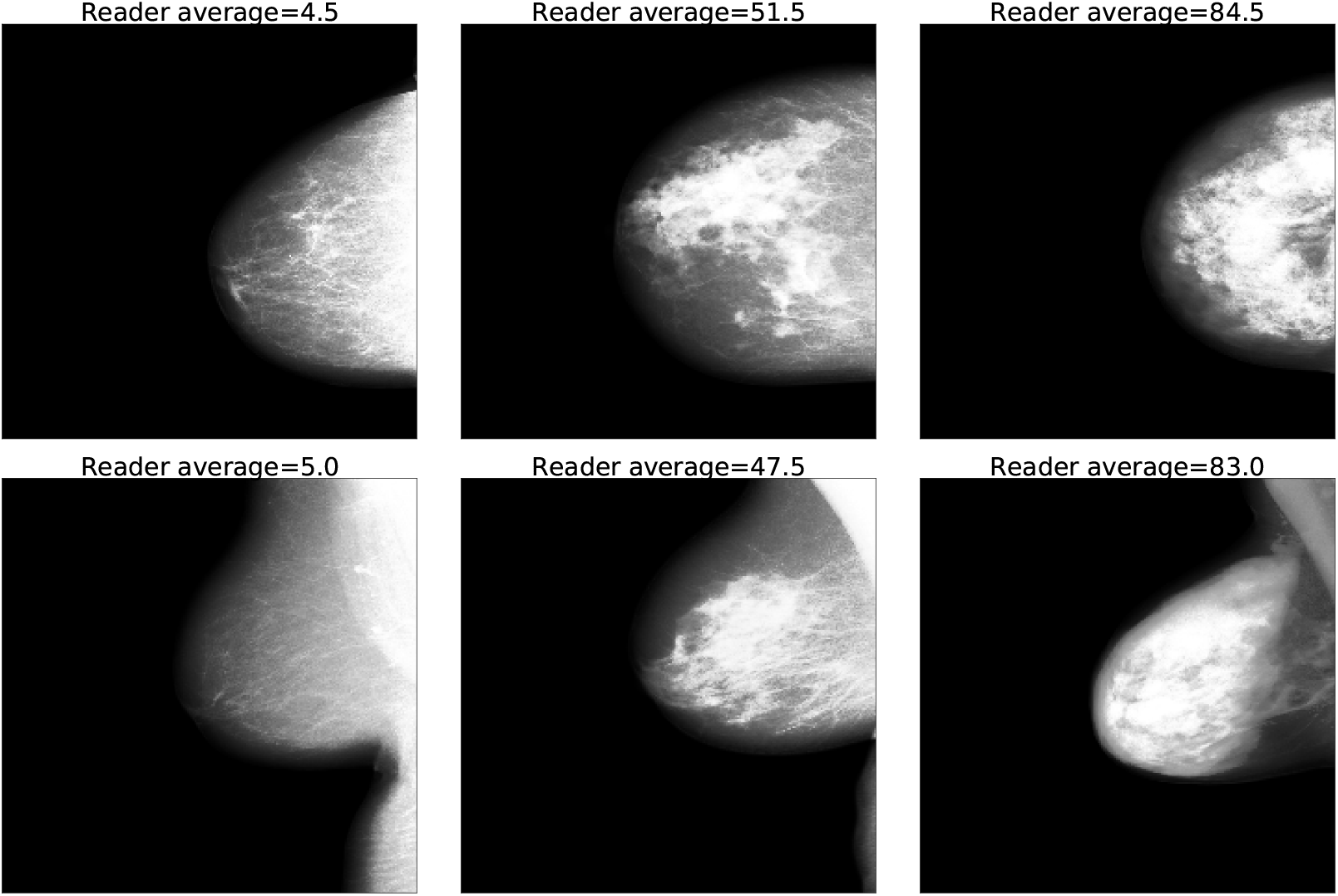
Images of low density (left), middle density (middle) and high density (right) images as defined by the average percentage density. These are the images after pre-processing (see Algorithm 1 in Section 3.1 for details). The top row are CC images and the bottom MLO.

We partition the data-set into training, validation and testing sets by woman, so that all the available views for a women are in the same partition. All the women from a previous case-control set,^4^ discussed below, are placed in the testing set. Otherwise the rest of the training, validation and testing sets are chosen at random from the PROCAS dataset. In total we have 33,011 women in the training set, 3,649 women in the validation set and 2,697 women in the testing set.

In addition, to compare with a previous study,^15^ we have a second partition with 19,048 women in the training set, 769 women in the validation set and 19,844 women in the testing set. Similarly to the previous partition all women from the previous case-control set ^4^ are in the testing set.

We also investigated the log-ratios for developing cancer from a previously created subset of the data^4^ called the priors which consist of women who did not have a cancer detected when their screen was taken but subsequently went onto develop a cancer. All this data was held in the test set and not used for training or validation.

These cancer risk predictions (on the priors) are found by taking the breast density estimates and splitting them into quintiles. Three control (no breast cancer) mammograms are matched with one breast cancer prior mammogram by matching on other known risk factors (age, BMI, hormone replacement therapy use, menopausal status and year of mammogram). This attempts to isolate breast density as a risk factor whilst controlling for potential confounding variables.

Therefore it allows for estimates of the ratios of probability of developing cancer for women with high breast density compared to low breast density. The log-ratio is calculated using conditional logistic regression on quintiles of the density. The higher the risk ratios at high density compared to low density the better the density model is at assessing risk. For further details of the approach see Astley et al.^4^

## 3 Prediction Methods

The objective is to take in a mammogram image and output an estimate of the density score of that image. The procedure we follow has four parts: 1) pre-processing stage; 2) using pre-trained deep learning models to extract features from the processed image; 3) mapping the features to a set of density scores; 4) using an ensemble approach to take the multiple scores and produce a final density estimate. Throughout this paper we will use “density mapping” to refer to the third step, where individual feature vectors are mapped to a density score and “ensemble prediction” to refer to the fourth step which takes those density estimates and combines them into a final ensemble prediction. In Figure 2 we show this process in a schematic format.

**Fig 2.**
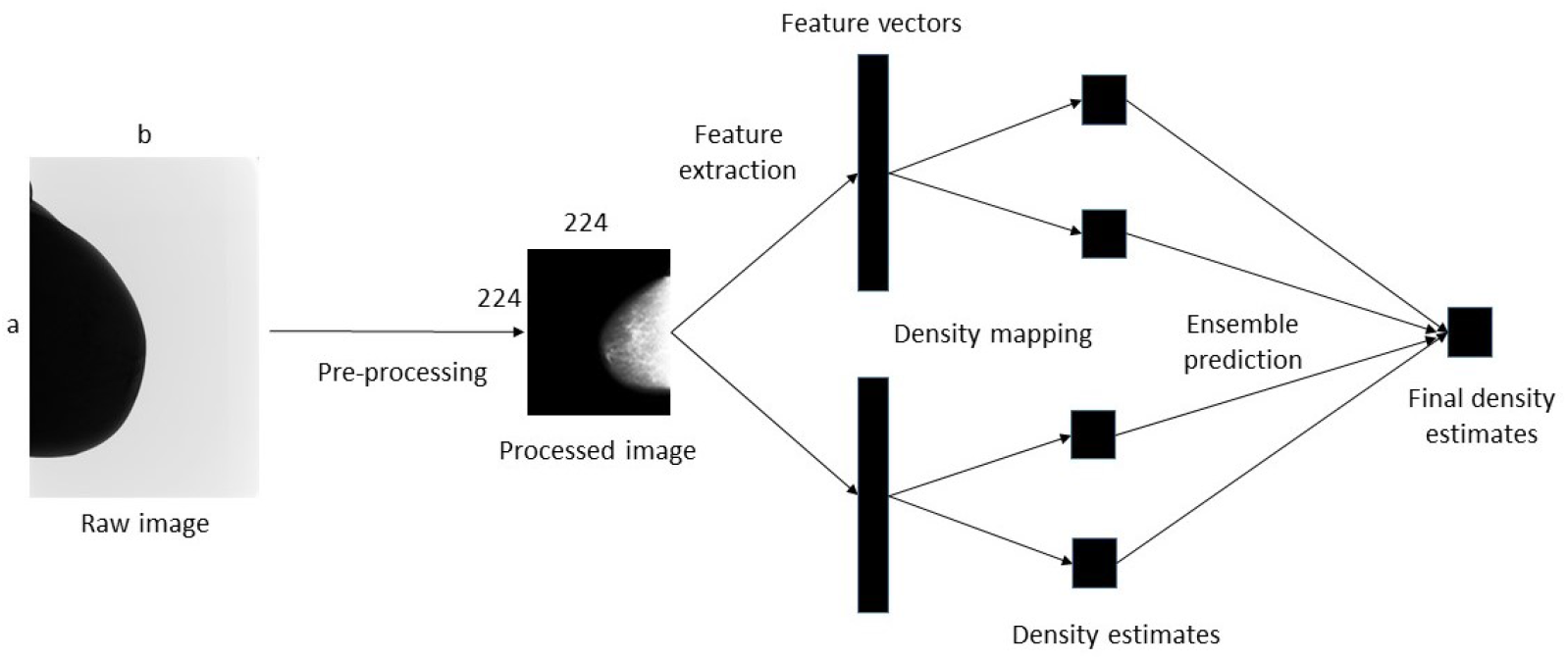
Illustration of our deep learning pipeline for producing final density estimations on mammograms. The input mammogram is processed to reduce the size down from (*a* × *b*) to (224 × 224) and increase the contrast. The processed image is fed into the feature extractors, we use two in this paper (ResNet^19^ and DenseNet^20^) but others could be included, which each produce a feature vector for that image. The density mapping, either a linear regression or MLP, is then applied to each feature vector to produce the separate density estimates. Finally the ensemble model is applied to convert the separate density estimates into one final prediction.

### 3.1 Pre-Processing

We pre-processed the images to size 224 × 224 (50, 176 input pixels) for processing by the feature extractors. The down-scaling process reduces the number of input pixels down to 1.1%, 0.68% and 0.22% of the original size for the small, medium and large images respectively (see Table 1 for the three image sizes).

We also enhance the contrast of the images to make it easier for the methods to extract information. In Algorithm 1 we lay out the steps we take to pre-process the images. We first rescale the image from its original size (see Table 1 for the three image sizes) down to 224 × 224 using cubic interpolation. We then clip all element values to 75% of the image maximum, subtract the minimum and divide through by the new maximum. The values are inverted and any image that is positioned on the left hand side is flipped horizontally. We perform histogram equalisation and normalise the image to contain values between 0 and 1. We show examples of these preprocessed images in Figure 1.

### 3.2 Feature Extraction

To perform the feature extraction part of our procedure we use two pre-trained deep networks: ResNet^19^ and DenseNet.^20^ Both were trained on the ILSVRC 2012 version of ImageNet,^9^ a large database of 1.2 million images across 1000 classes. ResNet and DenseNet are both popular in the literature, available as easily accessible models from PyTorch^21^ and produce modest sized feature vectors which keeps the computational requirement manageable. However, there are other potential feature extraction networks available that could be further investigated such as VGG^22^ or Inception.^23^

To extract features from the pre-processed images we remove the final fully connected classification layer from both networks which alters the output from 1,000 classes to 2,208 and 512 dimensional feature vectors for DenseNet and ResNet respectively. Details of our implementation is in Appendix A. We do not adjust the weights of the network or perform any form of fine-tuning.

#### Algorithm 1

Pre-processing

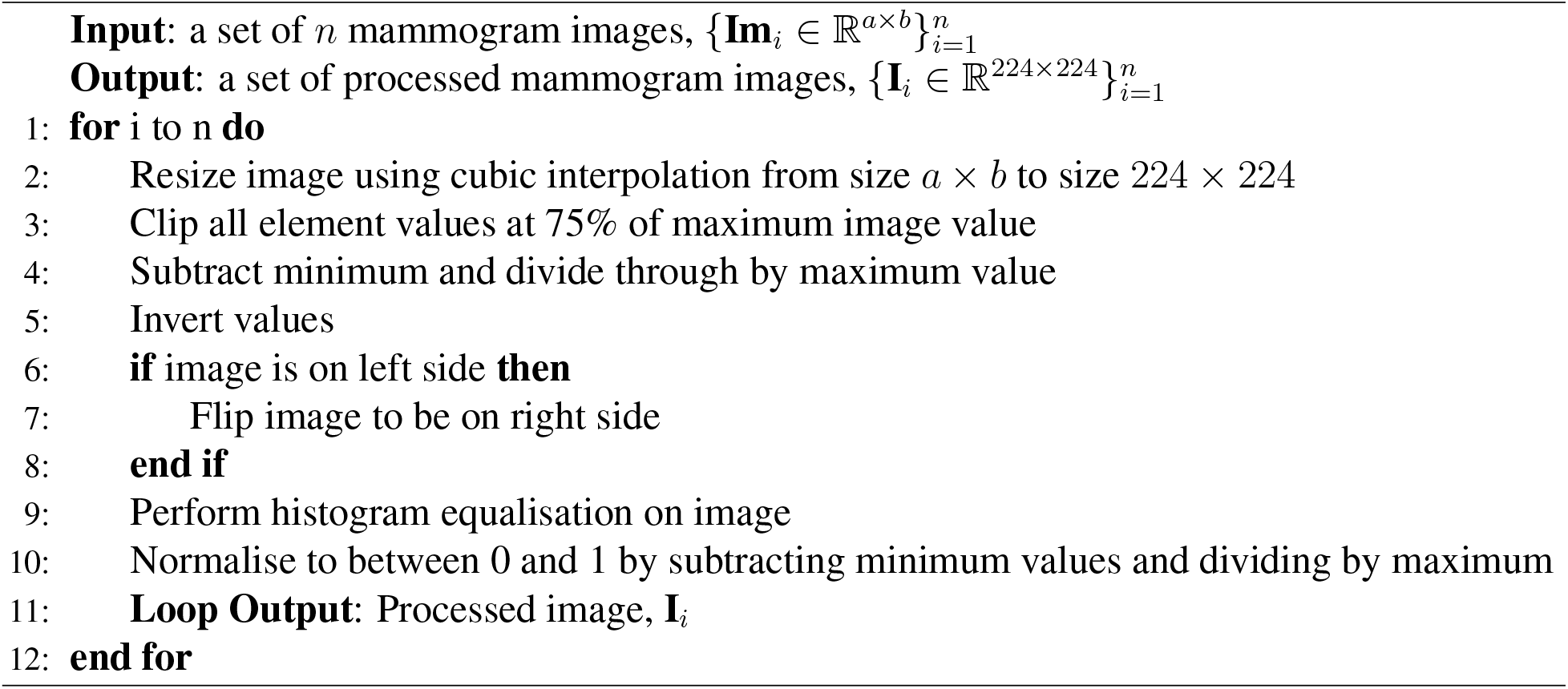

The training for the feature extractors was performed using ImageNet data which consists of natural images with three channels while our mammograms only have one channel. We therefore copy across the same image to make a repeated three channel tensor. Both networks require the inputs to be normalised across the channels to have means of [0.485, 0.456, 0.406] and standard deviations of [0.229, 0.224, 0.225] respectively. In Algorithm 2 we lay out the steps of our feature extraction method.

#### Algorithm 2

Feature Extraction Method

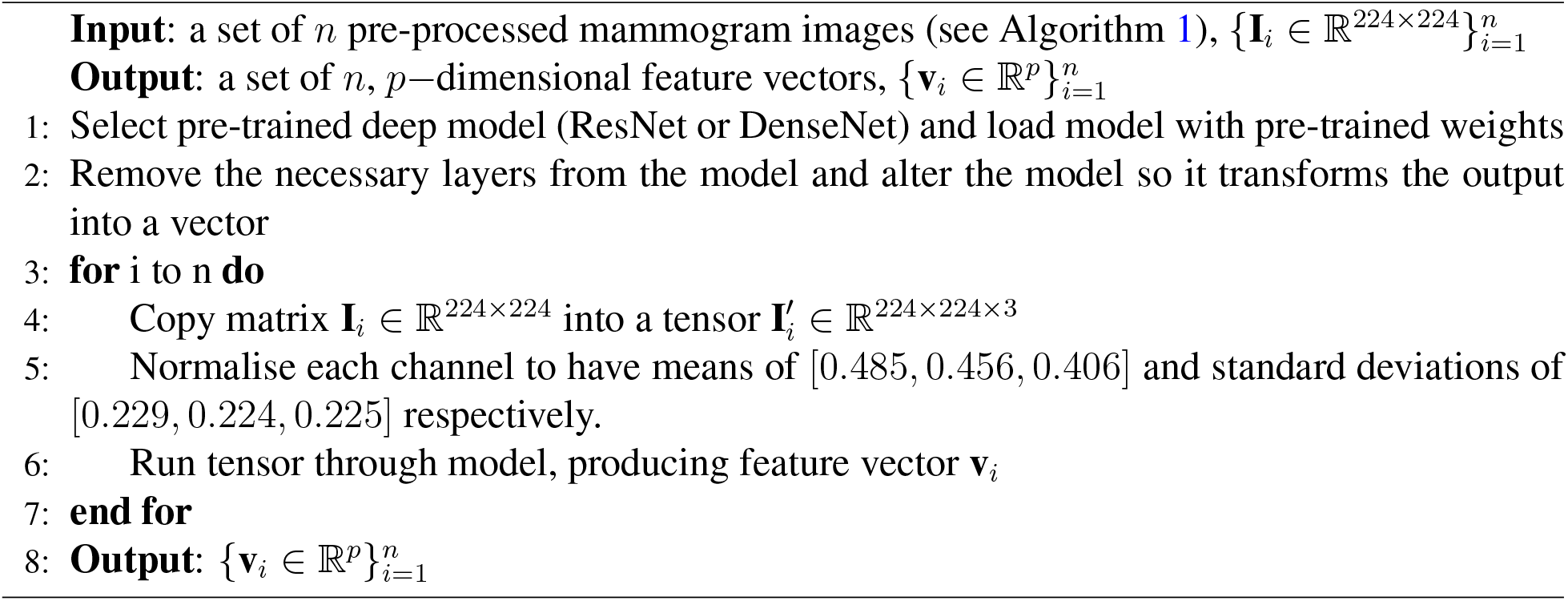

### 3.3 Density mapping

To map the deep feature vectors to produce a density estimation we utilise two methods: linear regression with regularisation and the application of multi-layer perceptrons (MLPs). The linear regression approach enables us to see how well a simple model, with one consistent solution, performs. An MLP can (in principle) map any function,^24^ and allows us to explore whether non-linear mappings are necessary.

A procedure for our linear regression method is shown in Algorithm 3. During training we add a bias term to the feature vectors from the training set, 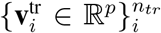, then stack them into a feature matrix, 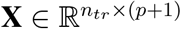. We utilise standard ridge regression, forming an objective function, 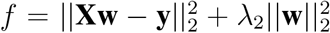, where 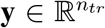 are the known labels and **w** ∈ ℝ^*p*+1^ are the weights. To find the optimal *λ*_2_ term we perform five-fold cross-validation on the training data, finding the value of *λ* which minimises the combined held-out error. We then retrain the weights on the entire training data-set using the optimal *λ*. To solve for **w** during both cross-validation and for the final optimal *λ* we use pseudo-inverse inversion: **w** = (**X**^*T*^ **X** + *λ***I**)^*−*1^**X**^*T*^ **y**.

#### Algorithm 3

Density Mapping: Linear regression training

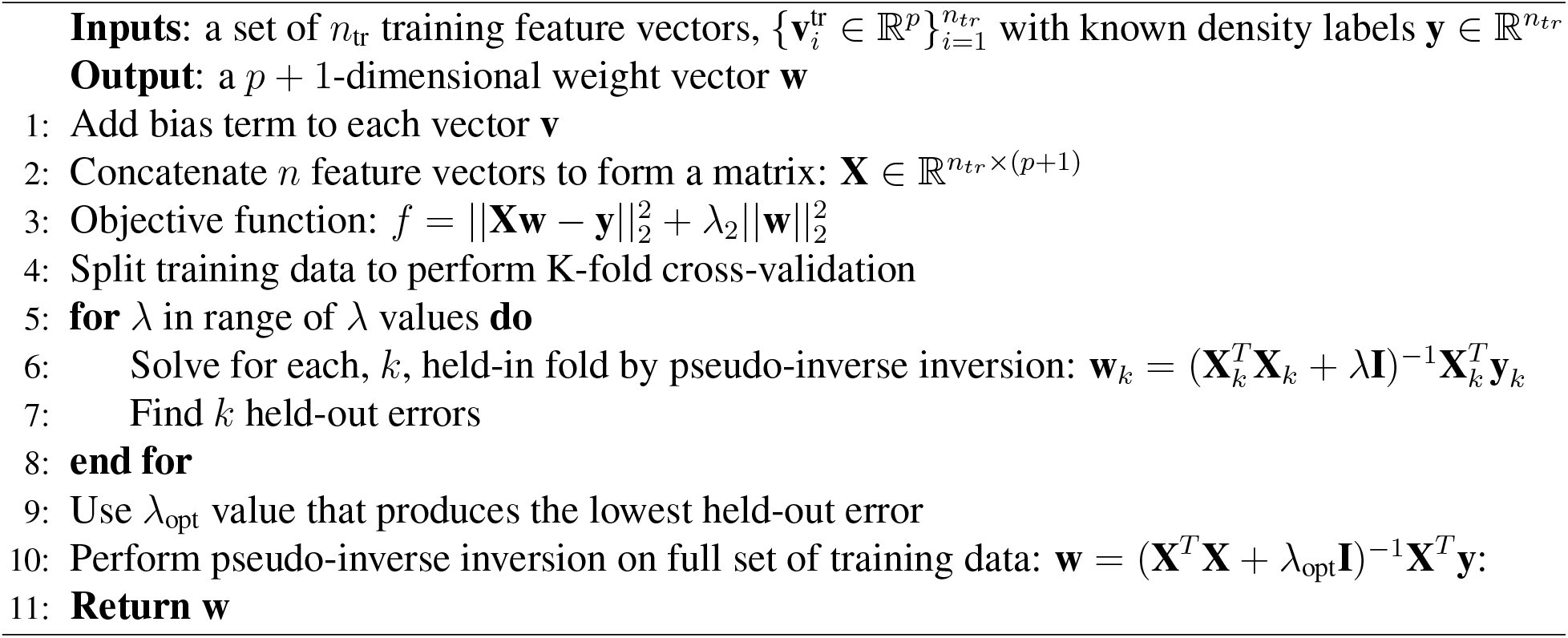

The linear regression approach assumes that the feature extractors are mapping the images onto a reasonably linear space where there is no need to consider non-linear correlations. As this assumption may not be correct, and it may be possible to improve performance with a more complex mapping we also consider the use of the multi-layer perceptron (MLP).

In Algorithm 4 we show the procedure we follow for applying the MLP to the data. We have the same input training vectors as for the linear regression. The details of the architecture and training are in Appendix A. In summary: we train the MLP on the same training partition as the linear regression, use three objective functions: *L*_1_, *L*_1*Smooth*_, mean squared error (*MSE*) and select the learning rates and training epochs from a trial and error approach until the training error converges reasonably smoothly to a steady state.

### 3.4 Ensemble Approach

The two deep feature extracting models along with a linear regression and the MLP models produce different predictions for each image. In addition, as will be discussed in Section 5.1, we can train on different labels, producing another set of different density mapping models. Ensemble methods tend to outperform individual models^25^ so if we can combine the individual predictions together we would expect to improve the model performance. Therefore there are 16 separate predictions: two feature extractors (ResNet and DenseNet) and four density mappings (linear regression and three MLPs trained with different objective functions). Those eight models can be either trained on averaged labels or individual labels (see Section 5.1 for details), giving the total of 16 separate sets of predictions.

#### Algorithm 4

Density Mapping: MLP

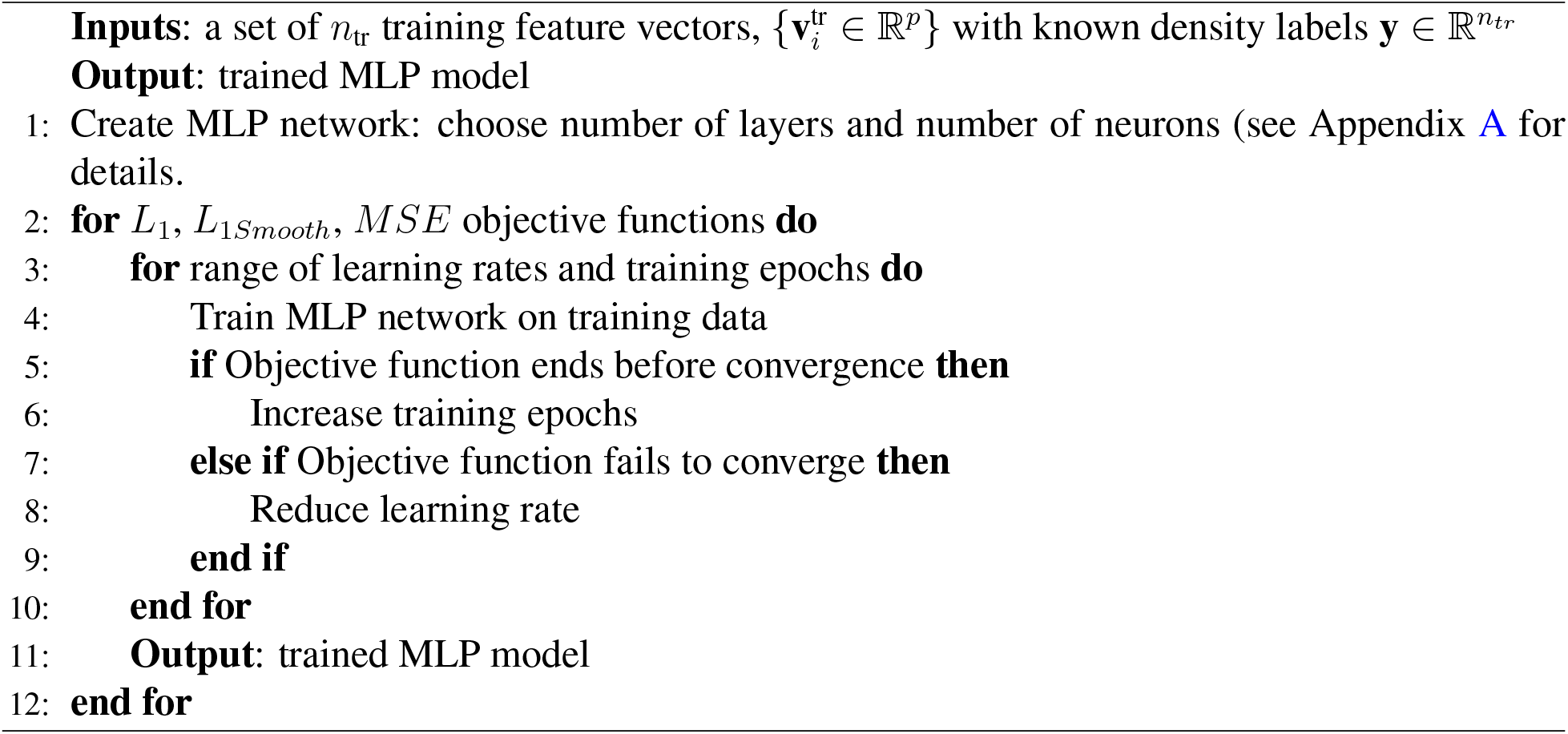

To produce our final ensemble prediction we start by splitting the training data into two sets - a larger one that each individual model is trained upon (see Algorithms 3 and 4) and a smaller one to train the ensemble on. We train the *m* individual models on the first training set and then apply each to the second training set to produce *n* predictions. We stack the predictions into a new training set which we then train a new MLP on to find a final model. To make predictions we run all *m* models on an image, produce the output for each one and make a final prediction by feeding the *m*-dimensional vector into our ensemble model. In Appendix A we provide details of the architecture and training procedure. There are many ensemble approaches available^26^ including simple methods such as averaging across individual results. We show just one approach that demonstrates that we can improve on individual results using an ensemble method.

#### Algorithm 5

Ensemble Method

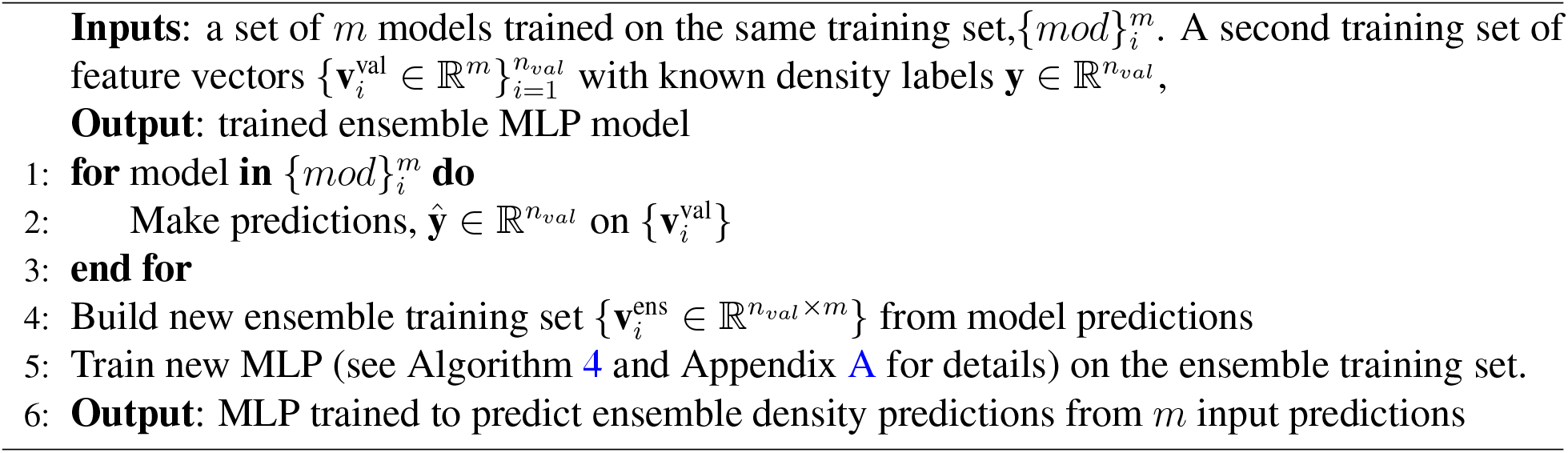

## 4 Assessing Predictive Performance

### 4.1 Metrics

To assess the predictive performance of our models we consider a range of metrics. The global measures we use to compare the quality of the VAS predictions with labels are Pearson correlation coefficient, root mean squared errors (RMSE), mean absolute error (MAE) and median absolute error (MedAE). We also show results of the risk ratios on the priors as discussed in Section 2.

### 4.2 Perfect Predictor Estimates

Due to label unreliability a “perfect” model which correctly estimates the VAS scores for all images will, if compared to the noisy labels, have a non-zero error. Therefore we need to produce an estimate of what metrics a “perfect” model would produce so that we can assess the quality of our approach.

We take the average of the two reader scores to be the “true” values and then assume that the actual reader scores are drawn from a Gaussian distribution around the real score. The Gaussian error distribution varies for different parts of the score distribution, with smaller average errors at both low and high densities, when considering averaged reader scores. We calculate the error distribution for small ranges of densities. We then use the “true” values, the averaged reader scores, and create a pair of modelled reader scores by adding Gaussian noise to the “true” value.

In detail: we take the average reader scores and call these our perfect modelled estimates. We bin the average reader scores into small bins (4% each) which provides a reasonable number of data-points to calculate the Gaussian parameters, except for above 80% where there is little data so we use the range 80-100% as one bin. The distributions approximate a Gaussian (see Figure 13 in appendix) but the Gaussian tails are longer than the real results, and there is the problem of modelled reader estimates below 0 and above 100. To correct for these two effects we resample from the distribution if the reader estimate is outside the 0-100 range or if the deviation of the sample from the mean is greater than a certain threshold *σ*_max_. These corrections make the modelled distribution a plausible match for the real data. To make the model as simple as possible the only parameter we alter is *σ*_max_. To check the model produces sensible estimates we compare the differences between the pairs of real reader estimates and the modelled pair differences. We can then adjust this tuneable parameter to equalise this comparison for the different metrics we consider. In this manner we can estimate the range of metrics that would occur for an optimal prediction method. We summarise our method in Algorithm 6.

We compare the four metrics (correlation, RMSE, MAE and MedAE) between the pair of modelled reader scores and match up the metrics between the pair of real reader scores. For example, we alter *σ*_max_ until the modelled pair of readers have the same RMSE as the real pair of readers. We do the same for the other metrics and record the optimal metrics from the lowest to the highest. We then show the range of these values when we perform the simulation.

## 5 Results

We split our results section into three subsections. In Section 5.1 we analyse the labels to gain insight into their reliability and how we need to train our models. Then in Section 5.2 we analyse the performance of our models compared to the ground truth as produced by radiologists labels of the images. Finally, in Section 5.3, we make a range of comparisons both between different versions of our models and between our models and previous work to try and gain further insight into our model performance.

### Algorithm 6

Perfect predictor model

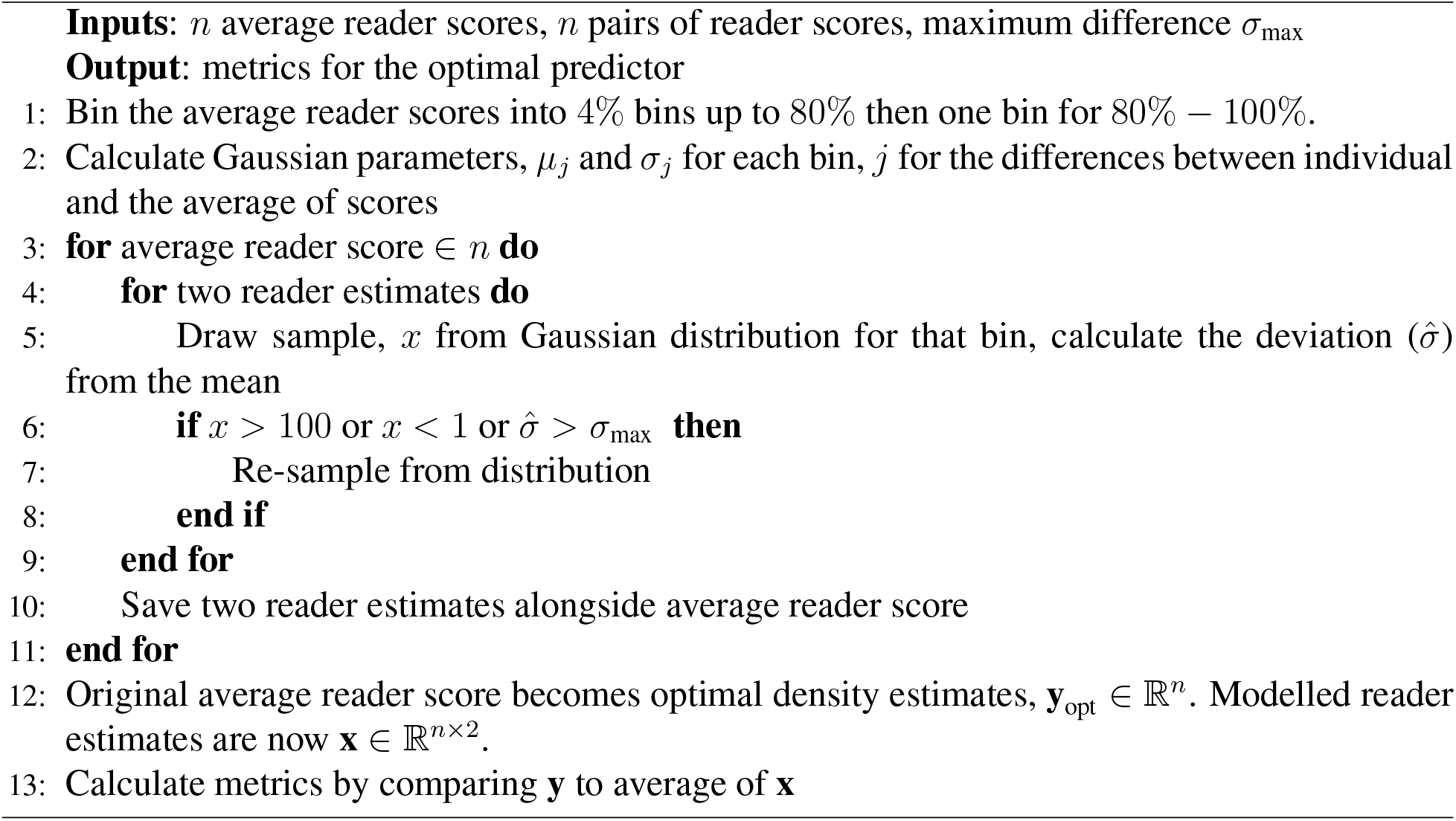

### 5.1 Reader Density Label Analysis

In Figure 3 we show the distribution of the density scores (left plot) and the distribution of the absolute differences between reader scores (right plot). For the density scores we show both the average of the two readers (*Averaged*) and the individual reader estimates (*Individual*). As there are twice as many individual scores as averaged scores we normalised the distributions to make them directly comparable. The distributions of density are highly skewed, with little data at high breast densities and relatively large amounts with a density of around 20. In addition, the averaged reader score tends to compress the distribution away from both low and high densities compared to the individual scores. The reader absolute differences (right plot) show a distribution with a relatively long tail, with many images having similar density scores but some with large differences.

**Fig 3.**
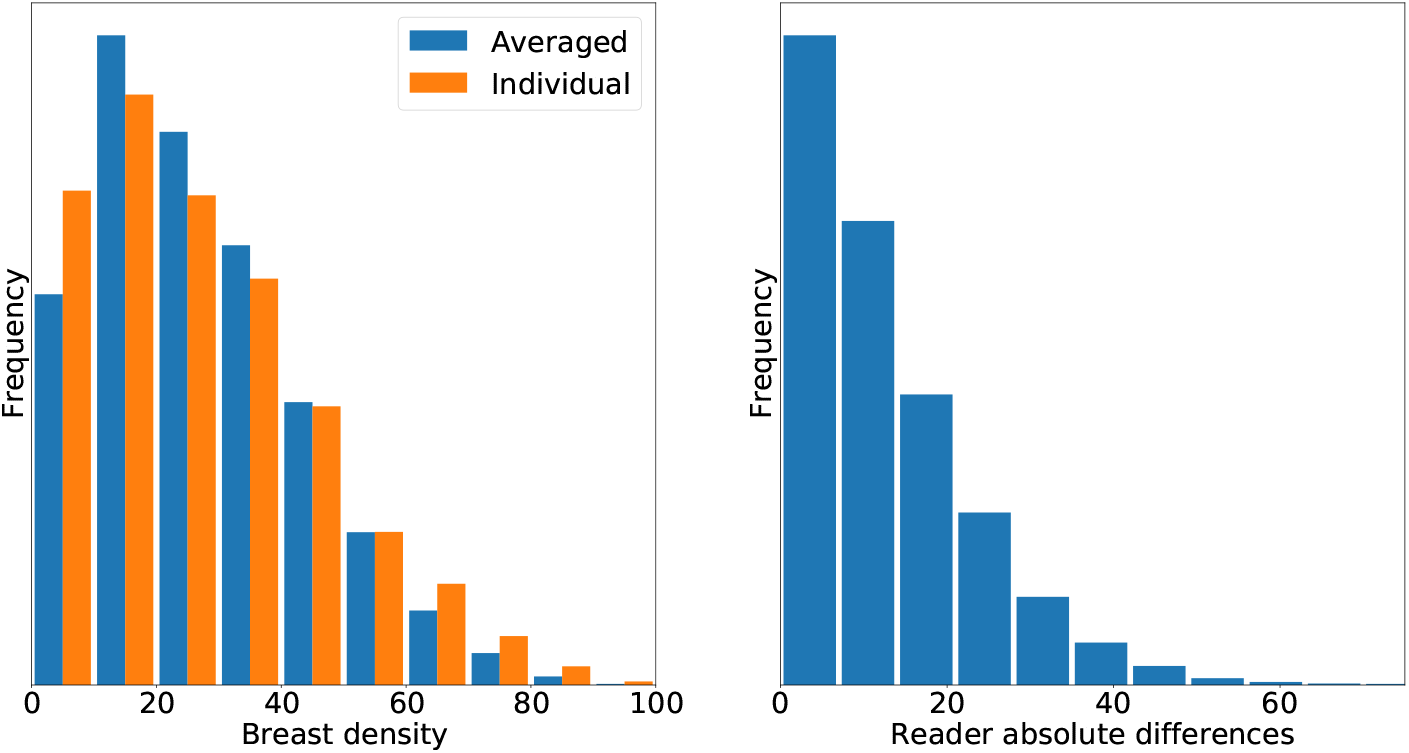
Left) Distributions of the averaged and individual VAS estimates. Right) Distribution of the absolute differences between reader scores.

In Figure 4 we show how RMSE between the pair of readers differs per decile of VAS scores for averaged and individual reader scores. The averaged and individual scores are used to define the deciles (bins) and the differences per decile then calculated using those images in each bin. In Table 6 in the appendix scores for mean absolute error and median absolute for the same decile are shown.

**Fig 4.**
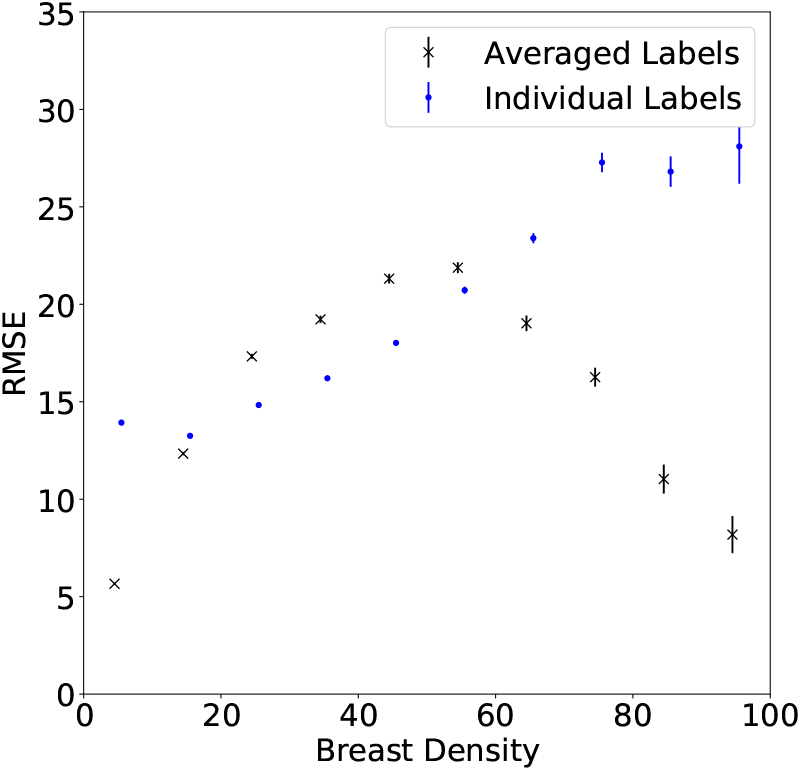
The RMSE per decile with the deciles determined either with averaged (crosses) or individual (dots) labels. The errorbars are at the 95% level estimated by bootstrapping with 1,000 repeats.

The variability of reader scores means that some of the images will be labelled inaccurately. To gain some intuition about the scale of this effect, in Figure 5 we show the distribution of the differences for the reader estimates using individual reader estimates as the scores. We bin the images into their deciles using the individual labels and then plot the distribution of the differences for each decile. The differences shown are between an individual VAS score and its pair, therefore we see skewed distributions.

**Fig 5.**
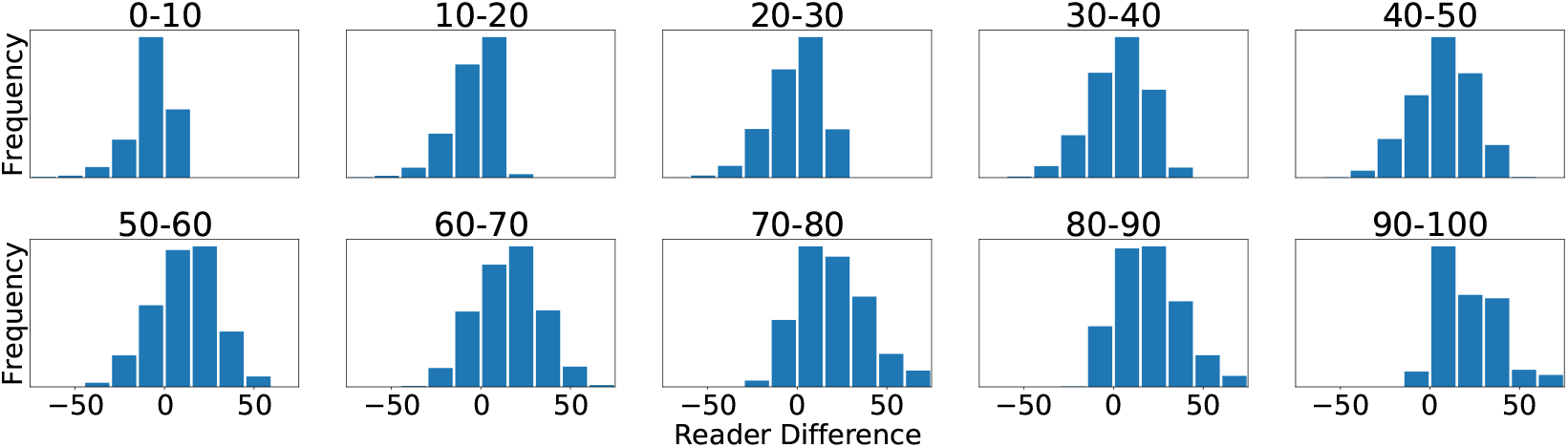
The distribution of the differences for each decile, considering the individual reader estimates. The decile is shown at the top, the reader difference axis is the same for all plots while the frequency varies depending on the number of images in each decile. The lower density scores tend to have tighter distributions with fewer large differences in label annotation.

In Section 4.2 we presented a simulation method for producing quantitative estimates of the errors for a perfect set of predictions. In Table 2 we show error measures between the modelled density scores and the reader estimates for correlation, root mean squared error (RMSE), mean absolute error (MAE) and the median absolute error (MedAE) for the entire data-set (*All data*) and for the test set. The ranges are by adjustments of *σ*_max_ (see Section 4.2 and Algorithm 6). There are further plots and analysis of these results in the appendices.

**Table 2.**
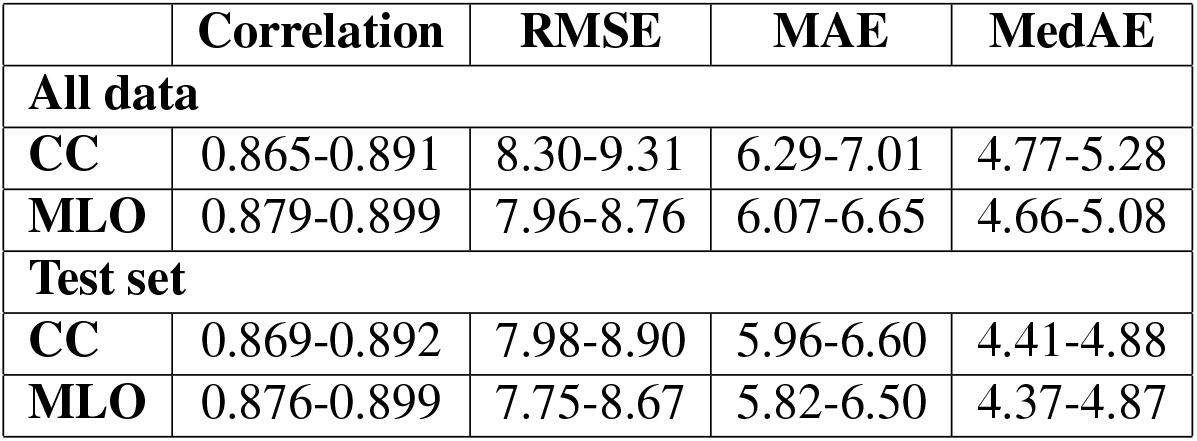
Expected metrics per image, for a model predicting the true VAS values if the assumptions we specify are correct. This can be seen as an estimate of a set of metrics we would find if we produced a highly accurate model. Correlation: Pearson correlation coefficient. RMSE: root mean squared error. MAE: mean absolute error. MedAE: median absolute value. The range is found by matching the metrics of the modelled pair of reader scores using *σ*_*max*_ for the four metrics. The best score (high correlation and low errors) is found when matching RMSE and the worst when matching the correlation.

|  | Correlation | RMSE | MAE | MedAE |
| --- | --- | --- | --- | --- |
| **All data** |  |  |  |  |
| **CC** | 0.865-0.891 | 8.30-9.31 | 6.29-7.01 | 4.77-5.28 |
| **MLO** | 0.879-0.899 | 7.96-8.76 | 6.07-6.65 | 4.66-5.08 |
| **Test set** |  |  |  |  |
| **CC** | 0.869-0.892 | 7.98-8.90 | 5.96-6.60 | 4.41-4.88 |
| **MLO** | 0.876-0.899 | 7.75-8.67 | 5.82-6.50 | 4.37-4.87 |

### 5.2 Model Predictions

In Figure 6 we show plots of our ensemble CC image density estimates against the reader average. The left plot is a direct comparison of prediction against reader average and on the right is a Bland-Altman plot of the difference between prediction and reader averages against the average of the two. Plots per woman are shown in the appendix in Figure 18 and show a similar pattern, with a smaller variation.

**Fig 6.**
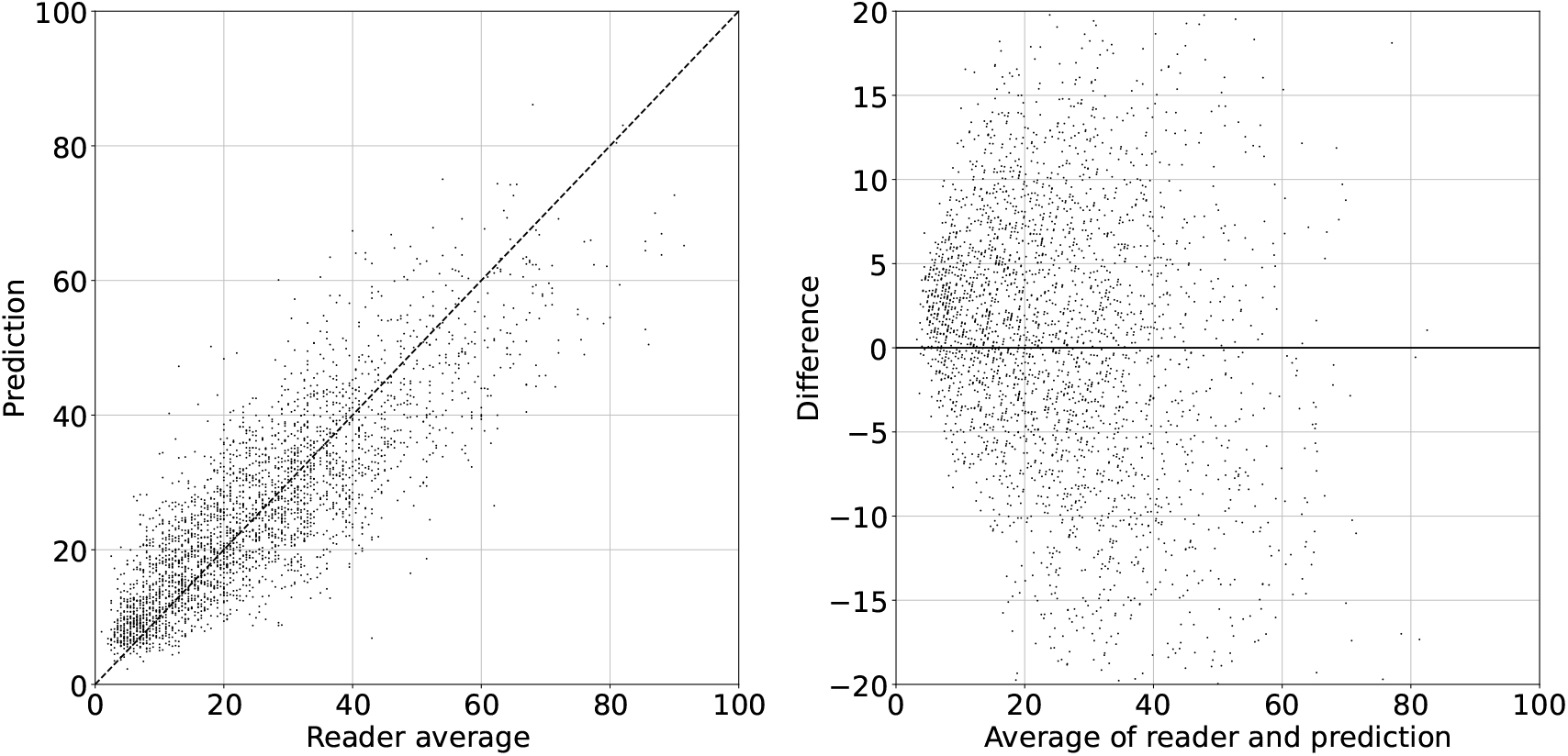
Density prediction of the CC images by the ensemble model compared to the averaged reader image estimates. Left) Direct comparison between the ensemble model prediction and the reader average score. The dashed line represents equal predictions and reader average scores. Right) Bland-Altman plot of the difference between the ensemble model prediction and the reader average score against the average of the two.

In Table 3 we show the four metrics produced by comparing our predictions against the averaged labels for the CC images. The equivalent results for the MLO images are in Table 7 in Appendix B. All the results are shown against the averaged reader scores of the test set. The *Label* columns refers to which label was used when training the models on the training data.

**Table 3.** Comparison metrics between our models and the average labelled data for the CC images.

| Model | Label | Obj. func. | Corr | RMSE | MAE | Median |
| --- | --- | --- | --- | --- | --- | --- |
| ResNet | Average | LinReg | $0.80 \pm 0.01$ | $9.50 \pm 0.21$ | $7.39 \pm 0.16$ | $5.97 \pm 0.20$ |
| | | L1 | $0.82 \pm 0.01$ | $9.34 \pm 0.22$ | $7.08 \pm 0.16$ | $5.48 \pm 0.18$ |
| | | L1Smooth | $0.82 \pm 0.01$ | $9.30 \pm 0.22$ | $7.05 \pm 0.16$ | $5.48 \pm 0.18$ |
| | | MSE | $0.82 \pm 0.01$ | $9.20 \pm 0.21$ | $7.10 \pm 0.16$ | $5.69 \pm 0.16$ |
| | Individual | LinReg | $0.80 \pm 0.01$ | $9.49 \pm 0.21$ | $7.38 \pm 0.16$ | $6.00 \pm 0.20$ |
| | | L1 | $0.82 \pm 0.01$ | $9.37 \pm 0.21$ | $7.04 \pm 0.16$ | $5.34 \pm 0.19$ |
| | | L1Smooth | $0.82 \pm 0.01$ | $9.38 \pm 0.22$ | $7.06 \pm 0.16$ | $5.28 \pm 0.20$ |
| | | MSE | $0.82 \pm 0.01$ | $9.13 \pm 0.21$ | $7.00 \pm 0.15$ | $5.52 \pm 0.18$ |
| DenseNet | Average | LinReg | $0.82 \pm 0.01$ | $9.12 \pm 0.21$ | $7.08 \pm 0.15$ | $5.72 \pm 0.16$ |
| | | L1 | $0.83 \pm 0.01$ | $9.03 \pm 0.22$ | $6.87 \pm 0.15$ | $5.38 \pm 0.22$ |
| | | L1Smooth | $0.83 \pm 0.01$ | $9.04 \pm 0.21$ | $6.88 \pm 0.15$ | $5.38 \pm 0.19$ |
| | | MSE | $0.83 \pm 0.01$ | $8.99 \pm 0.21$ | $6.94 \pm 0.15$ | $5.56 \pm 0.18$ |
| | Individual | LinReg | $0.82 \pm 0.01$ | $9.12 \pm 0.20$ | $7.08 \pm 0.15$ | $5.71 \pm 0.16$ |
| | | L1 | $0.83 \pm 0.01$ | $9.05 \pm 0.22$ | $6.77 \pm 0.16$ | $5.19 \pm 0.2$ |
| | | L1Smooth | $0.83 \pm 0.01$ | $9.05 \pm 0.23$ | $6.77 \pm 0.16$ | $5.11 \pm 0.18$ |
| | | MSE | $0.83 \pm 0.01$ | $9.01 \pm 0.21$ | $6.88 \pm 0.15$ | $5.39 \pm 0.18$ |
| Ensemble | | L1 | $0.84 \pm 0.01$ | $8.79 \pm 0.21$ | $6.66 \pm 0.15$ | $5.17 \pm 0.17$ |

Figure 7 shows differences in labels per decile (left plot) and prediction errors per decile (right plot). The label differences are slightly different than Figure 4 (but there are no differences in trends) as those were on the entire data-set and these on the test set. The images are binned either by their averaged labels (crosses) or individual labels (dots). The errorbars are the 95% confidence intervals found by performing 1,000 sets of bootstrapping. The prediction errors are taken as the difference between the ensemble predictions and averaged labels, the individual labels are used to bin the images, not to estimate the accuracy of the model.

**Fig 7.**
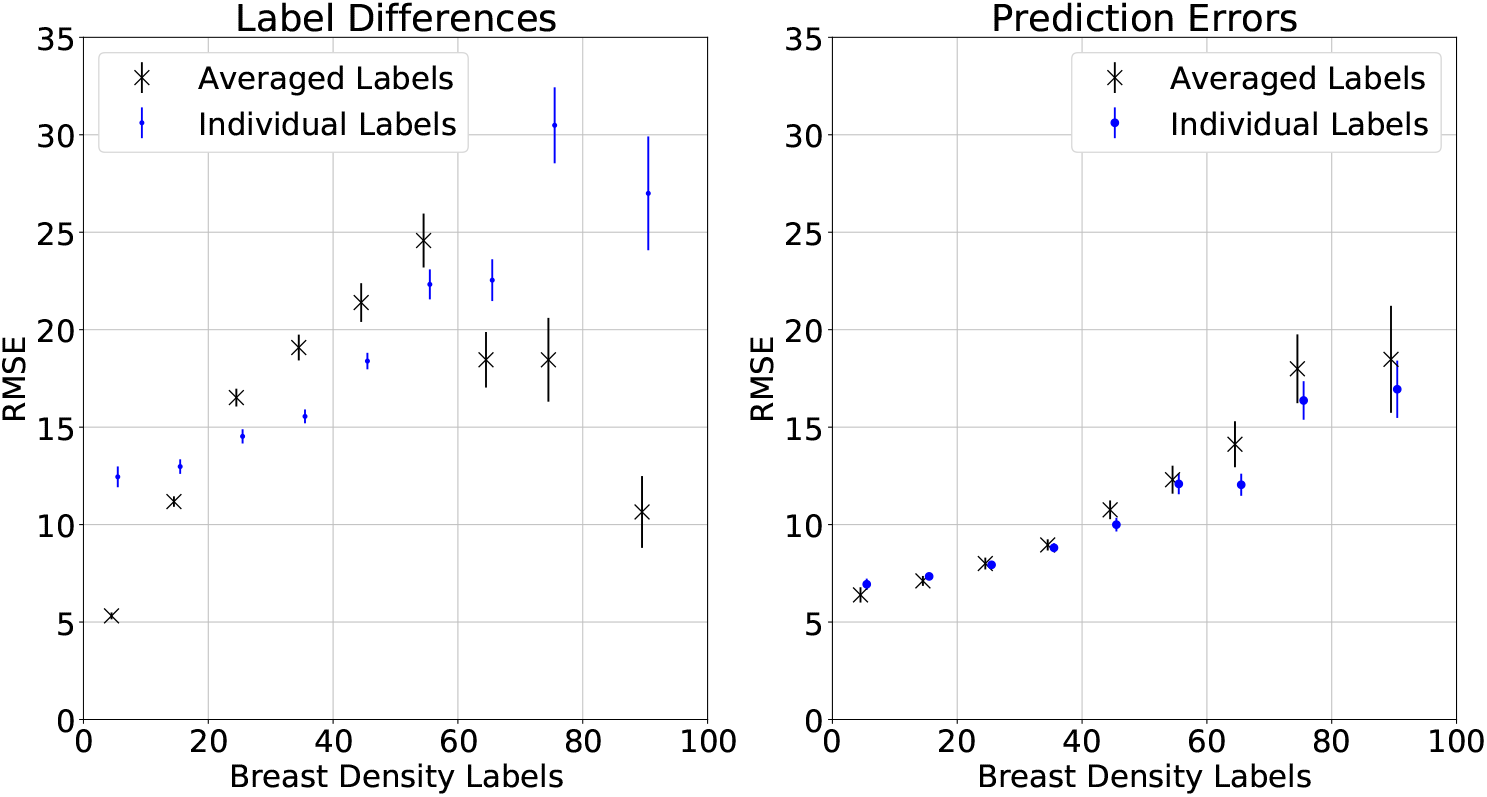
The changes in RMSE per decile for the labels and ensemble predictions, with deciles defined by averaged and individual labels. The error bars are the 95% confidence intervals found by bootstrapping 1,000 times. Left) Reader differences as measured by RMSE with data placed into deciles from averaged labels (black crosses) and individual labels (blue dots), respectively. Right) Ensemble prediction RMSE per decile with data placed into the decile using the average label (black crosses) and individual labels (blue dots)).

In Figure 8 we show the results of cancer risk predictions for the same 16 model variants and ensemble predictor as in Table 3. The odds ratios (OR) are in comparison to the first quintile. The most relevant is Q5 which shows the OR of the highest density women compared to the lowest density women.

**Fig 8.**
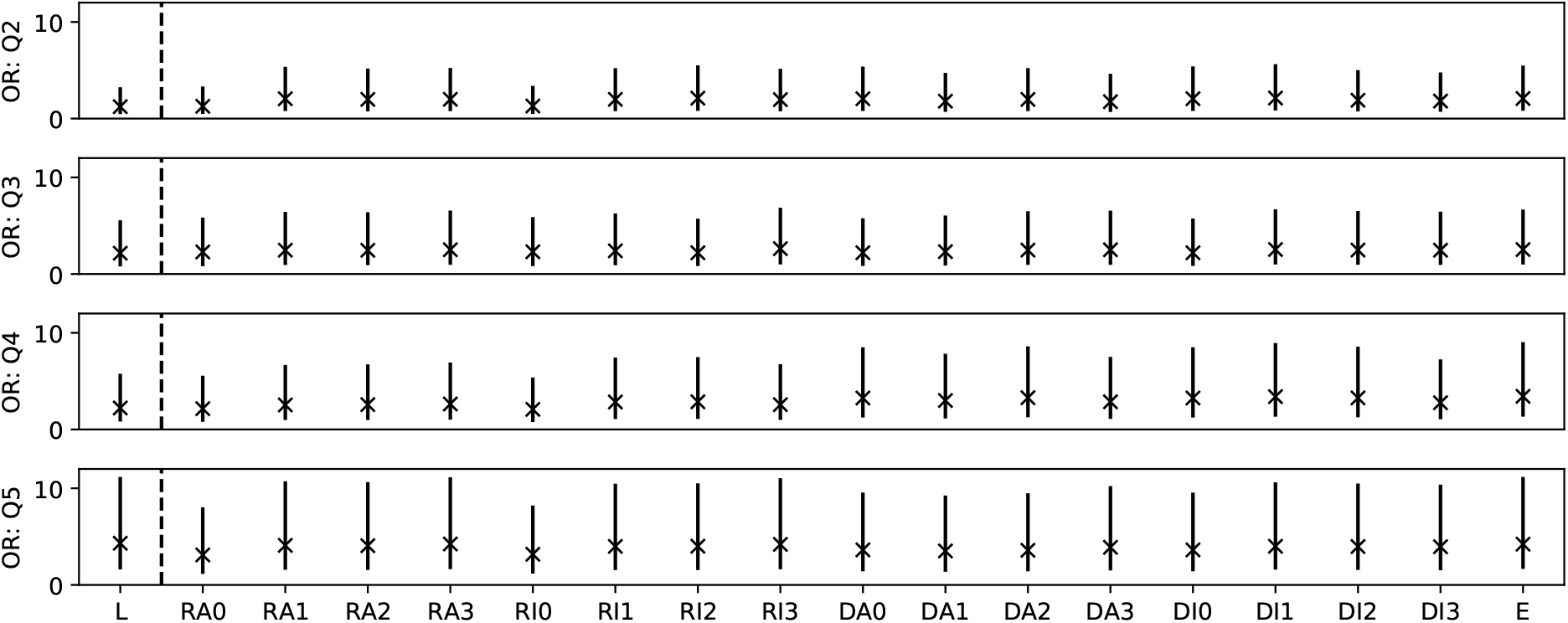
Plots of the odds ratios calculated using the model predicted scores with uncertainties found via bootstrapping. *OR: Q2,Q3,Q4,Q5* shows the odds ratios for the second, third, fourth and fifth quintile respectively. *L* shows the odds ratios found from the labels. *R/D* represent ResNet or DenseNet, *A* represents the model trained on the averaged labels, 0,1,2,3 represent linear regression, the L1, L1Smooth and MSE objective functions respectively. For example, *RI2* is the ResNet feature extractor trained on the individual labels with a L1Smooth objective function. *E* is the ensemble predictor.

### 5.3 Model Comparisons

We make a comparison between: models trained on averaged and individual labels; models with different feature extractors (ResNet and DenseNet); models with density mapping from linear regression and MLPs. In addition we compare our predictions to those of a variant of a previous method trained on this data, *pVAS*.^15^

In Figure 9 we show a comparison between model predictions trained on the averaged labels and the individual labels for the DenseNet MLP method with L1 objective function and in Table 4 (*Comparison 1*) we show metrics for the differences between these predictions.

**Fig 9.**
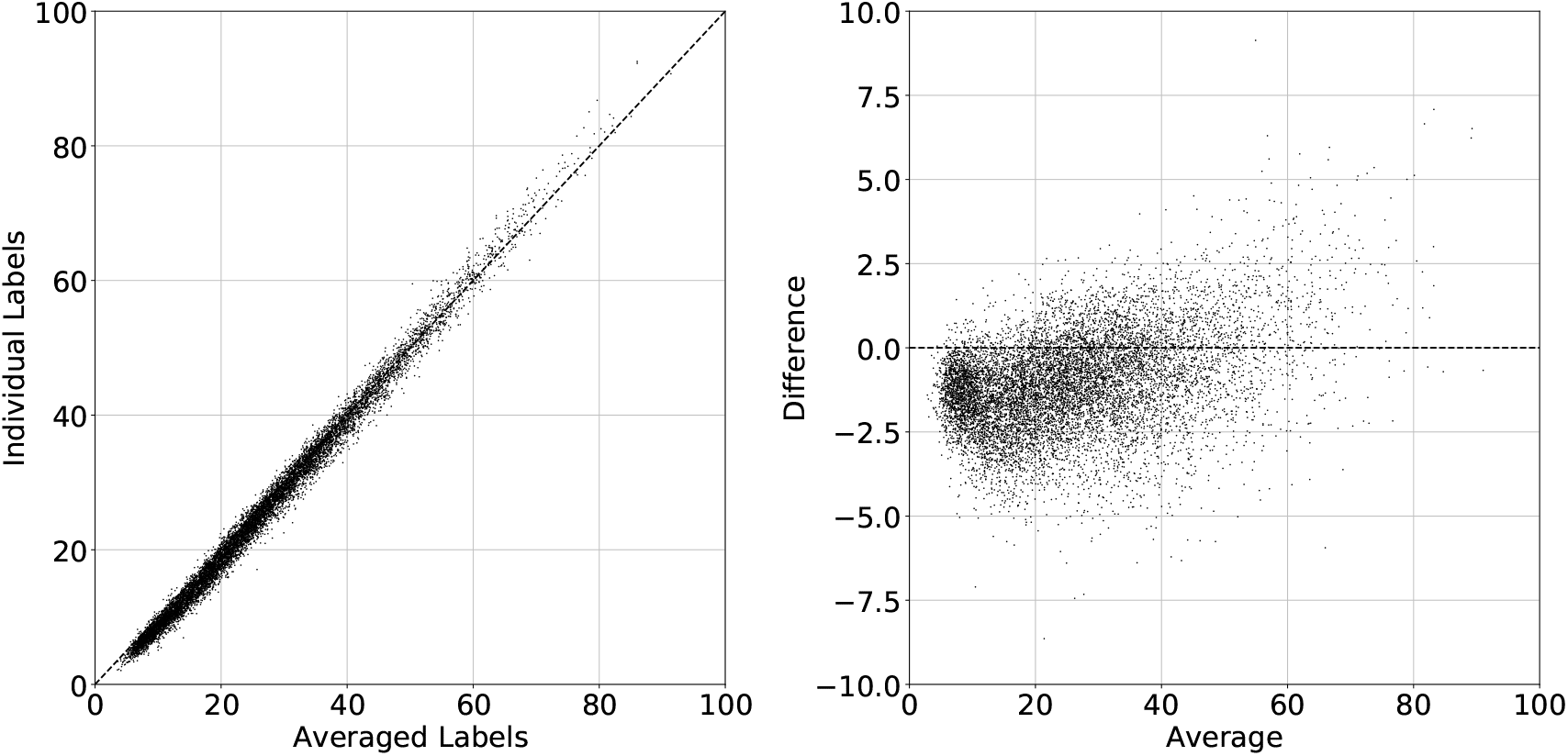
Plots showing the similarities and differences between image predictions made by models trained on individual labels and averaged labels. Left) Predictions from training on individual labels against predictions from training on the averaged labels. Right) Bland-Altman plots of the same data.

**Table 4.** Comparison metrics between the predictions made between our density mapping models. We show standard metrics with the addition of the RMSE per quintile (labelled **Q1**-**Q5**), with quintiles defined as the average of the two predictions. **AvQ** is the mean of the RMSEs per quintile. Comparison 1 is between models trained on averaged and individual labels. Comparison 2 is between MLPs trained on densenet and ResNet extracted features. Comparison 3 is between an MLP and linear regression model trained on the DenseNet extracted features. Comparison 4 is between our ensemble model trained on both individual and averaged labels and *pVAS* from a previous study.^15^ The test set for this comparison is different than in the rest of this paper.

| Name | Corr | RMSE | MAE | MedAE | AvQ | Q1 | Q2 | Q3 | Q4 | Q5 |
| --- | --- | --- | --- | --- | --- | --- | --- | --- | --- | --- |
| **Comparison 1** | 0.996 | 1.80 | 1.46 | 1.28 | 2.33 | 1.90 | 1.70 | 1.69 | 2.29 | 4.05 |
| **Comparison 2** | 0.945 | 4.69 | 3.55 | 2.72 | 6.54 | 3.27 | 4.9 | 6.25 | 8.44 | 9.82 |
| **Comparison 3** | 0.983 | 2.60 | 2.04 | 1.68 | 3.86 | 2.66 | 2.32 | 2.97 | 4.35 | 7.02 |
| **Comparison 4** | 0.924 | 5.74 | 4.40 | 3.53 | 7.89 | 4.63 | 5.80 | 7.00 | 8.85 | 13.19 |

In Figure 10 we show a comparison between model predictions made using the feature extractors ResNet and DenseNet, both with MLPs trained on the feature vectors. These are from training on individual labels and with the L1 objective function and is labelled *Comparison 2* in Table 4. The differences are larger than those trained on the individual and averaged labels (the axes of the Bland-Altman plot are twice the size of Figure 9) but they also appear to be random in character, there do not appear to be much in the way of systematic variation.

**Fig 10.**
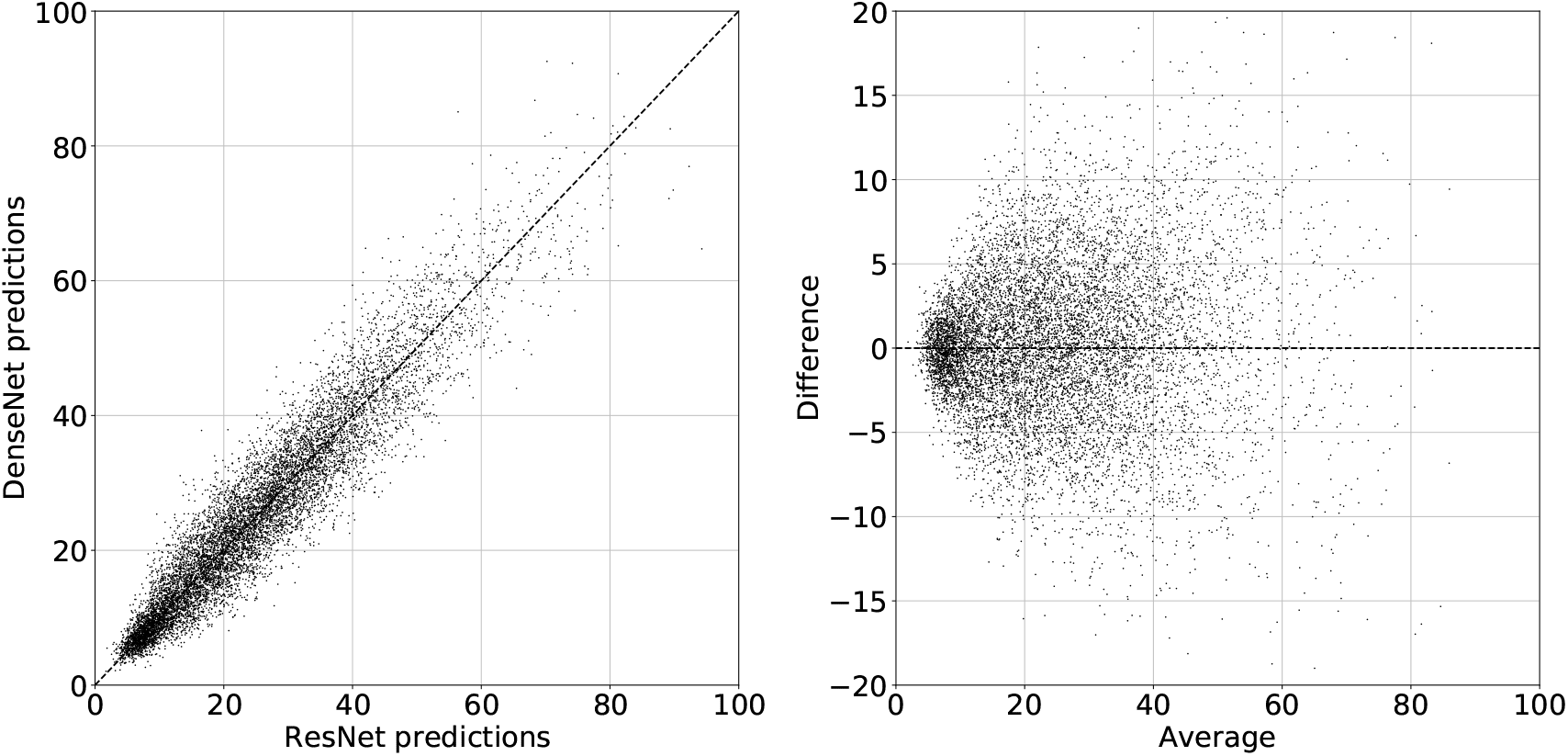
Differences in image prediction between MLPs trained on the DenseNet and ResNet feature vectors. Left) Predictions made using DenseNet against predictions made using ResNet. Right) Bland-Altman plots of the difference between predictions made by DenseNet and ResNet versus their average.

We show a comparison between model predictions made using the linear regression and MLP density estimators, both using the DenseNet feature extractor, in Figure 11. These are for average labels and L1 objective function for the MLP and is labelled *Comparison 3* in Table 4.

**Fig 11.**
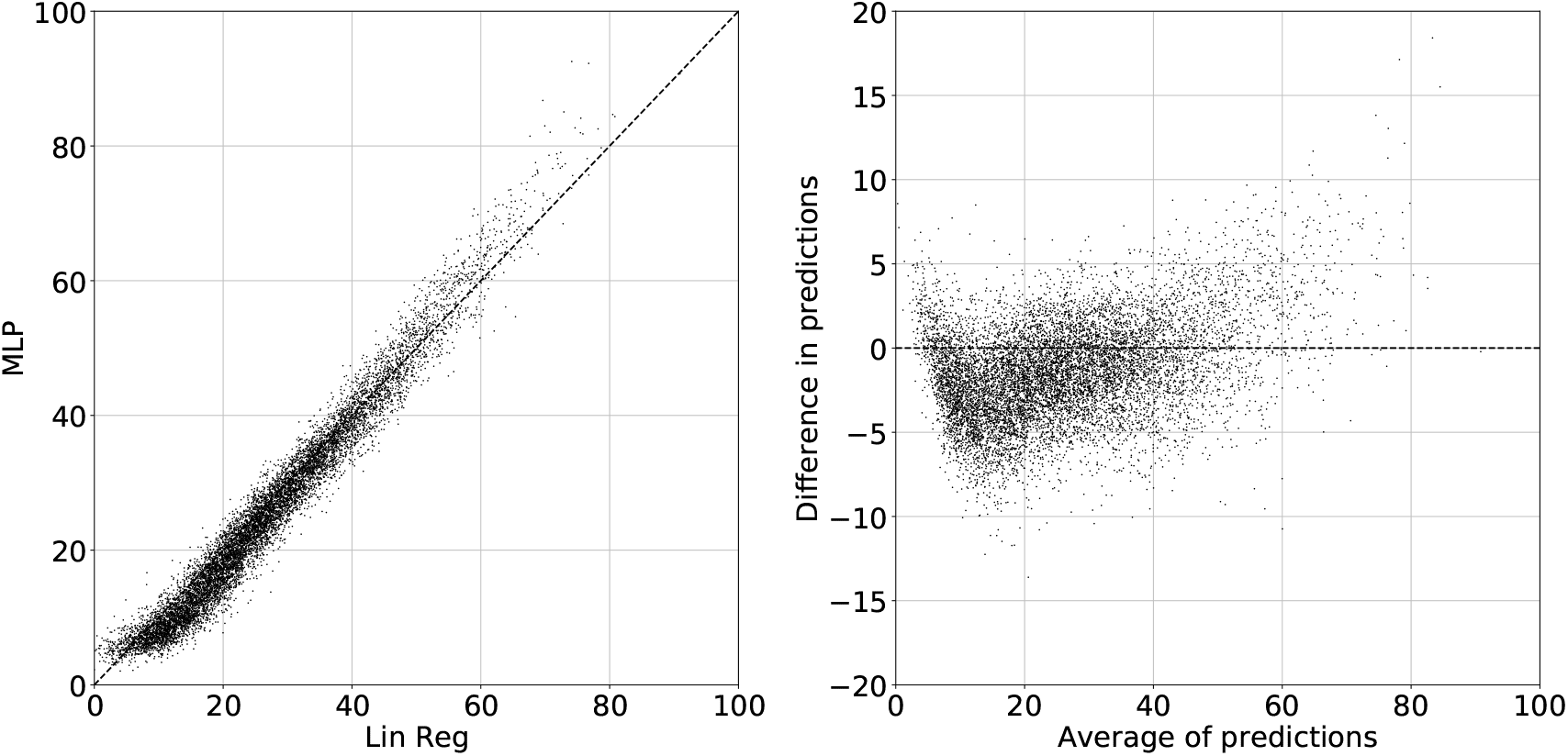
Image predictions made using DenseNet extracted features with a MLP and linear regression as the density mapping functions. Left) Direct comparison. Right) Bland-Altman plots of the difference between predictions made by MLP and linear regression versus their average.

In Figure 12 we show a comparison between previous work,^15^ labelled *pVAS*, and the final ensemble predictions produced in this paper. The test set is different to that for the results shown in the rest of the paper so that it coincides with the test set used by the *pVAS* model.

**Fig 12.**
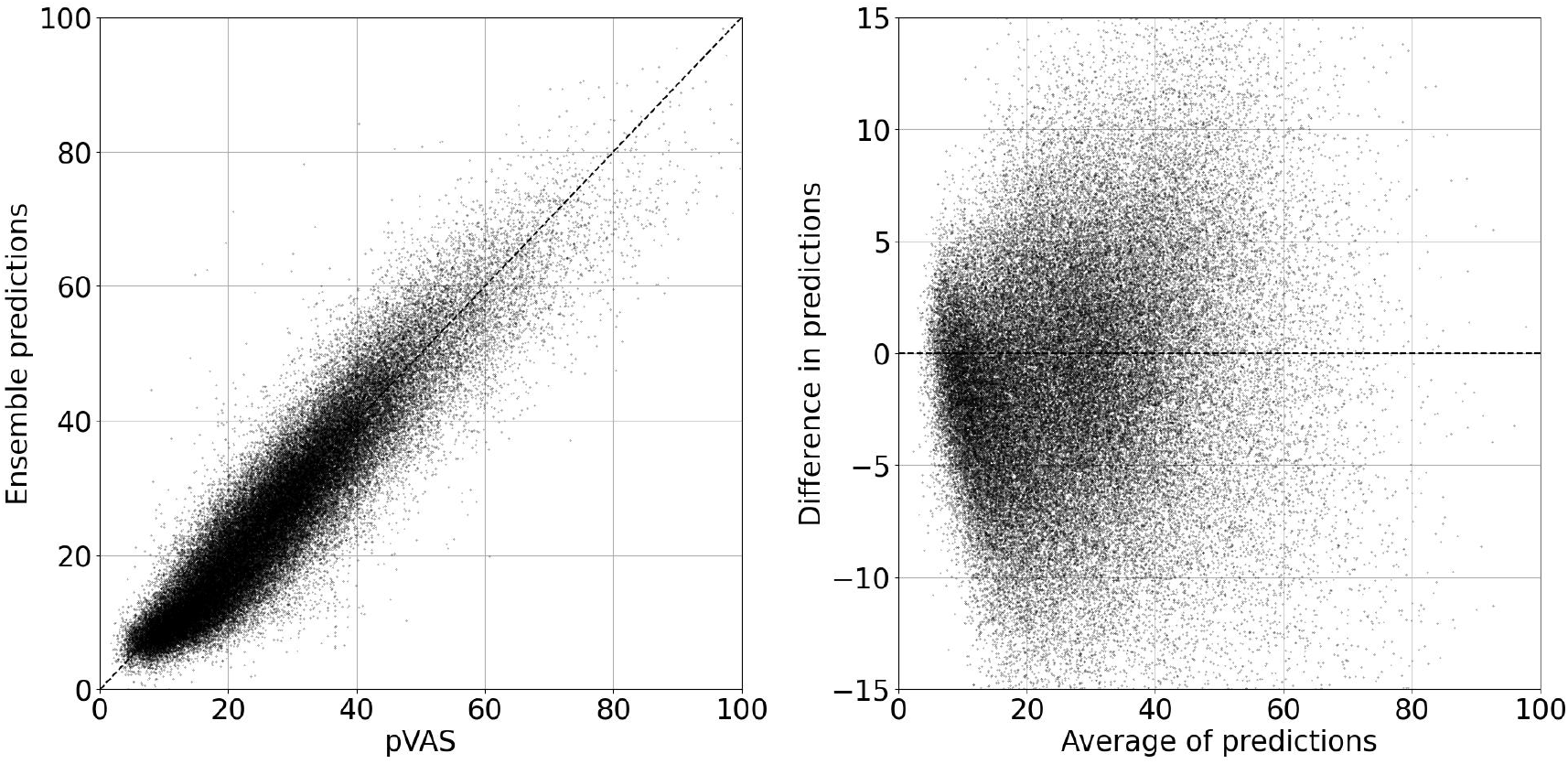
A comparison between the ensemble predictions using our methods and *pVAS*.^15^ Left) Direct comparison. Right) Bland-Altman plot.

Table 5 shows metrics for the *pVAS* estimates and our ensemble estimates. We also show these results as *Comparison 4* in Table 4.

**Table 5.** Metrics of predictions made by our ensemble method and predictions made by *pVAS*^15^ per image. The results are all the images, both CC and MLO. These results are from the second test set partition, discussed in Section 2. Uncertainties are at the 95% confidence level found via bootstrapping with 1000 repeats.

|  | **Correlation** | **RMSE** | **MAE** | **MedAE** |
| --- | --- | --- | --- | --- |
| *pVAS* versus our method | $0.924 \pm 0.001$ | $5.74 \pm 0.04$ | $4.40 \pm 0.03$ | $3.53 \pm 0.03$ |
| Our method versus labels | $0.839 \pm 0.003$ | $9.00 \pm 0.05$ | $6.92 \pm 0.04$ | $5.49 \pm 0.05$ |
| *pVAS* versus labels | $0.809 \pm 0.003$ | $9.78 \pm 0.05$ | $7.66 \pm 0.04$ | $6.32 \pm 0.05$ |

## 6 Discussion

Breast density, as measured on a visual analogue scale by experienced radiologists, shows a strong realtionship with breast cancer risk.^4^ We designed and implemented a frame-work based on pre-trained deep networks to find a mapping between a mammogram and its associated breast density. The core part of the system is the feature extractor which is taken from deep networks trained on ImageNet.^9^

Reader density scores are known to be variable.^27, 28^ One consequence is that we are training our models on data with inter-reader variability and the second is that if we were to produce a highly accurate predictor, when we compared it to the labels, it would look like it produced inaccurate predictions. We analysed the labels, in Section 5.1, to gain insight into how this variability affects our conclusions.

An important consideration is how the variability changes with the density scores. The average of the two reader estimates (Figure 4 and Table 6) appear to show significantly smaller differences between the readers at both lower and higher densities with a maximum difference in the middle. For example, in the 50-60 decile we have a mean difference of 18.0 and in the 0-10 decile a mean difference of 4.5. This might imply that the readers are more consistent at the two extremes and produce more variable results in the middle. However, if we look at the individual reader estimates the average differences between the readers tend to increase with increasing density across the deciles.

**Table 6.** The fraction of density scores (*Number Fraction*) and the average (both mean and median) absolute differences between reader estimates per decile. These are all shown for the deciles defined either by averaged reader scores (*Averaged*) or individual reader scores (*Individual*)

| Decile | Number Fraction |  | Mean Differences |  | Median Differences |  |
| --- | --- | --- | --- | --- | --- | --- |
|  | Averaged | Individual | Averaged | Individual | Averaged | Individual |
| **1-10** | 0.151 | 0.197 | 4.5 | 9.3 | 4.0 | 5.0 |
| **11-20** | 0.251 | 0.228 | 9.9 | 9.7 | 8.0 | 7.0 |
| **21-30** | 0.214 | 0.189 | 13.9 | 11.7 | 12.0 | 10.0 |
| **31-40** | 0.170 | 0.157 | 15.1 | 13.1 | 13.0 | 12.0 |
| **41-50** | 0.109 | 0.108 | 16.8 | 14.8 | 14.0 | 13.0 |
| **51-60** | 0.059 | 0.059 | 18.0 | 17.2 | 16.0 | 16.0 |
| **61-70** | 0.029 | 0.039 | 15.1 | 19.1 | 12.0 | 17.0 |
| **71-80** | 0.012 | 0.019 | 13.0 | 21.5 | 11.0 | 18.0 |
| **81-90** | 0.003 | 0.007 | 8.7 | 21.2 | 7.0 | 18.0 |
| **91-100** | 0.000 | 0.001 | 7.2 | 22.6 | 6.0 | 20.0 |
| **All** | 1 | 1 | 12.2 | 12.2 | 9.0 | 9.0 |

The reason the differences appear to peak in the middle of the density distribution and are lower at the two ends for the averaged reader estimates is likely to be at least partially a statistical artefact. If we consider the first decile (1-10) and take an example of an averaged reader score of 5.0. The maximum difference possible for this average result is one reader marking 1 and the other marking 9, with a difference of 8. Conversely the maximum possible difference for an average value of 50.0 is one reader marking 1 and the other marking 99, giving a difference of 98. Part of the reason that the average differences appear to be lower in the low and high densities is because, when considering the average scores, the results with the larger differences would not result in averages with low or high densities. There might be more label consistency at the low and higher ends of the density distribution, but it is hard to separate that out from this statistical artefact.

As the performance of our models is measured based upon the quality of the labels, if the label variability increases with higher density we would expect to see an apparent fall in model performance due to the increasing variability, rather than the failure of the model. In Figure 7 this is the effect we do see, an apparent reduction in performance of the model at higher density scores. It is likely that our models are more inaccurate at higher densities as we also see a large variability when comparing model predictions to each other at those higher densities (Table 4) but it is a smaller effect than if we simply measure the differences in predictions to the labels. These effects are further discussed in the appendix.

The errors between our density predictions (Table 3) and the labels fall close to the range of the errors we see with a modelled optimal estimator (Table 2) which can perfectly predict the density of an image. There is still space for improvement in the quality of the predictions, which may be best achieved by adapting the feature extractors to better represent mammography data either through fine-tuning or with other approaches. However, there is a limit above which it will become difficult to assess if the improved models are able to perform better as they approach the metrics shown for the simulated optimal model.

We compared our models both to a previous method, *pVAS*,^15^ and to variants of our method. We find considerable similarities between all of our models and to *pVAS*. This sort of similarity implies that we are finding true structure in the data. There is greater divergence of prediction at higher VAS scores, although it is still fairly small considering the uncertainty in the labels and the small amount of data at those higher densities.

There is little difference in results when training on averaged or individual labels (Table 3) and this is an interesting finding considering the low consistency of the pairs of reader estimates. We might expect there to be a significant improvement when training on the averaged labels because they might be expected to reduce some of the noise in data. The fact that we do not see this suggests that the models are able to effectively extract a true density related signal. The ensemble predictor produces a small improvement in performance giving slightly lower errors and higher correlations than the individual density mapping models.

The linear regression produced reasonable accuracy implying the feature extractors are producing a mapping to a fairly linear subspace, however, there is clearly some non-linearity required as the MLPs do perform better (Table 3). DenseNet performs better than ResNet, which may be due to the fact that the DenseNet version performs better on ImageNet than the specific ResNet version used (see Appendix A for details). Alternatively, it may be due to the larger subspace size of the DenseNet model.

The model predictors (Figure 8) are all comparable with the VAS labels in terms of risk prediction. While there is variation between the models in terms of the odds ratios found these are all within the uncertainty bounds and we cannot make any strong statements about the quality of the predictive models compared to one another.

This usage of transfer learning allows us to leverage the long training times and large computational power of other research groups. There is a debate over whether transfer learning or learning from scratch are more appropriate.^10^ One answer is that transfer learning approaches like the one we have demonstrated here is far quicker to implement than the requirement of designing and then training networks from scratch. If results from a transfer learning approach are poor then it then may be necessary to pursue other approaches.

A potential issue with a transfer learning approach in medical imaging is that the models used tend to be trained on unrelated images. It might be expected that this would result in features being extracted that are unrelated to the medical domain. However, we have shown that reasonably accurate results can be obtained using these features, even when using only a linear mapping. Perhaps with some domain adaptation these results would improve further.

We reduced the size of the images to 224 × 224 both to match the size of images the models were originally trained on and to reduce computational requirements. If we can utilise smaller images and achieve good results it is a significant advantage in terms of the computer power and time required. Saving on computational time is a major advantage, one benefit is that it allows researchers and groups without access to large computing resources to perform analysis. It also means that multiple different runs of algorithms can be performed and multiple other facets of the data can be investigated.

We also did not: preserve the aspect ratios of the images, do anything about the fact that there are three different image sizes which are then distorted by differing amounts or crop the images to focus on the breast. Yet we still achieve accurate predictions, further research is required to investigate whether results could be improved by correcting these issues, or if they do not adversely effect the quality of the models. We also do not utilise the three-channel nature of the transferred network, something that might enable more predictive capacity to be extracted from the pre-trained models.

Overall our method produces predictive accuracy close to the maximum that can be assessed with the ground truth labels we have access to. We do so using a method which requires modest computer resources both in terms of memory and time. In particular, once the feature extractors have been run the computational requirements of the density predictors are very low, especially for the linear regression. This can enable both much faster training but also training on small dataset or subsets of the larger data-set. We also demonstrate the, perhaps, surprising ability of deep learning models trained on a different image domain to produce good performance on this medical data-set.

There is a direct benefit of accurate prediction of breast density in that it can be used to produce information to medical practitioners or in research projects where there is no access to radiologists. Another potential value is as an input to other automated models. Density provides considerable information which might enable models that are trying to predict cancers, perform segmentation or other tasks might be able to utilise to produce improved performance on their specific problem.

## 7 Conclusions

In this paper we have demonstrated that using a transfer learning approach with deep features results in accurate breast density predictions. This approach is computationally fast and cheap which can enable more analysis to be done and smaller data-sets to be used. However, the deep feature models were trained on a non-medical data-set which would imply that the features extracted could be considerably improved. If we can train deep models on medical imaging data then we might expect to see improvements in performance when those models are used as a transfer learning model across a wide range of medical imaging applications.

We have demonstrated the issues associated with data where readers are variable in predictions. Finding ways to reduce the variability of the labels would enable us to train on more reliable data and to better assess which models are performing better than others. If we could improve the quality of the labels it would also mean we could more systematically investigate what measures could improve performance.

## Data Availability

Data is not currently available outside of the University of Manchester.

## Appendix A: Model Details

### A.1 Feature Extraction

We use two networks for the feature extraction: ResNet^19^ and DenseNet,^20^ specifically we use the “resnet18” and “densenet161” architectures downloaded with pre-trained weights from the PyTorch^21^ Torchvision repositories. For both we replace the final classification layer (“fc” in Resnet and “classifier” in DenseNet) with an identify matrix. We ran the images through the two networks with a NVIDIA Quadro P400 2 GB GPU, a process that takes around 10 and 50 seconds per 100 images for ResNet and DenseNet respectively. In total we have around 160,000 images and the total run time is around 4.5 and 22 hours for ResNet and DenseNet respectively. Once this stage is completed there is no need to repeat as the feature vectors remain the same and can be used in any required permutation.

### A.2 MLP Density Mapping

Our MLP is small and simple with *p* (512 for ResNet and 2208 for DenseNet) input neurons, followed by 200 neurons and a ReLU,^29^ then 300 neurons and a ReLU to the output. We trained them using the same NVIDIA Quadro P400 GPU on PyTorch^21^ with around 200 epochs, the Adam optimizer^30^ and a starting learning rate of 1 × 10^*−*5^ and one reduction in learning rate half way through to end at 2 ×10^*−*6^. Due to the small and simple nature of the MLP, multiple other training approaches would suffice, this one was found through trial and error checking that the training error was reducing as we would expect. We use three objective functions: L1, L1Smooth and the mean squared errors (MSE), all available as basic functions in PyTorch.

### A.3 Ensemble Prediction

The ensemble MLP is built using PyTorch^21^ with two hidden layers of size 5*p* and 10*p* respectively, where *p* is the number of density predictions fed into the system. We apply ReLU to both hidden layers and trained using the Adam optimizer.

## Appendix B: Extra results

In Figure 13 we show the distribution of the differences between individual label scores for the bins we use to make our simulated predictions. The bin centres are noted on the top of each plot.

**Fig 13.**
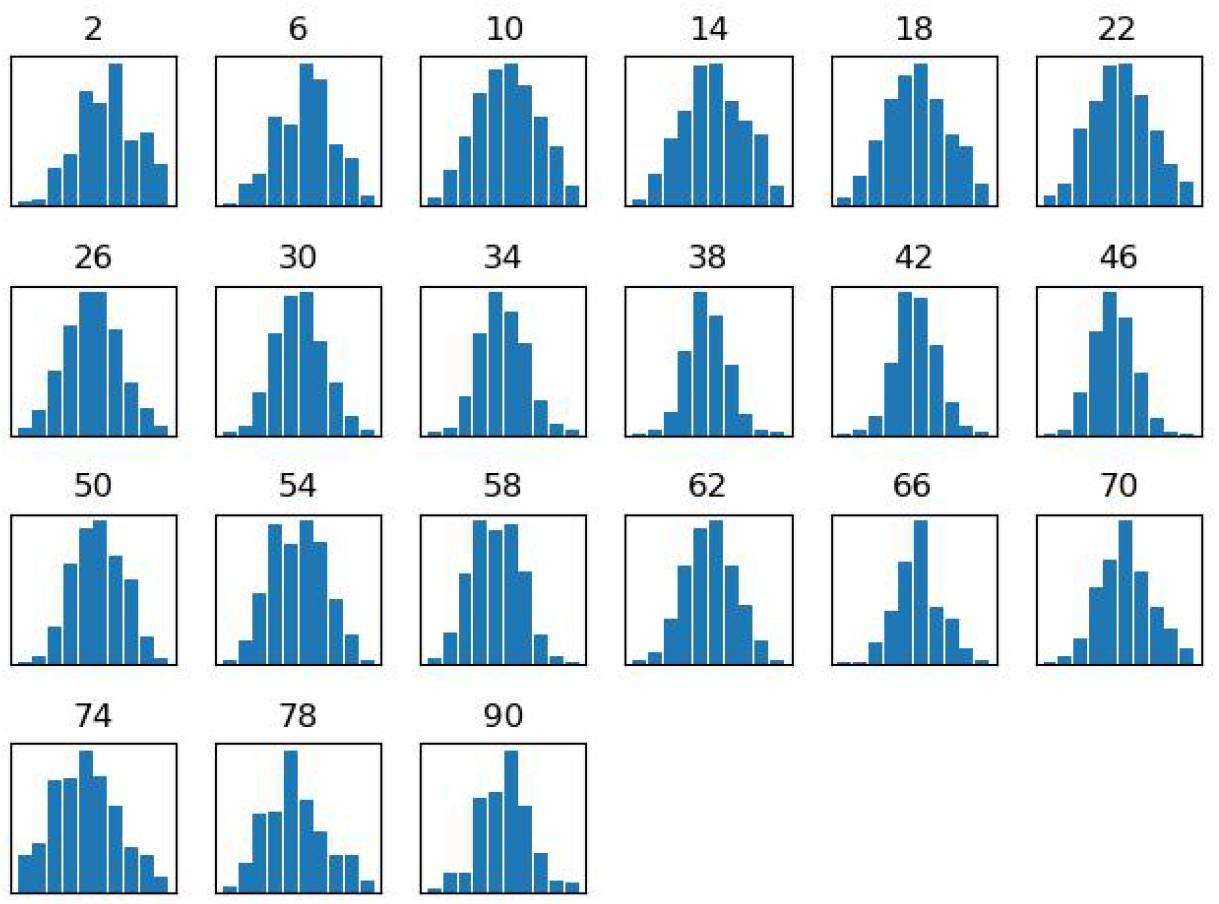
Distribution of the differences within bins. Bin centre are noted on the tops.

### B.1 Label Analysis

In Figure 14 we graphically demonstrate the challenge with having high reader uncertainty together with a skewed distribution. We plot all the binned individual reader scores per decile along with the distribution of the differences from the centre of the decile, these are the blue bars. We also plot the total number of individual labels above 80% as orange bars. In the bottom right two plots we show the entire distributions (left of the bottom right pair) and a zoomed in version (bottom right plot). The potential number of images which are falsely labelled as high density, may be comparable to the number of real high density images. This makes it difficult to confidently assess how well our models are performing at high VAS scores as we cannot assess whether a high VAS score is accurately labelled. If we were to perform over-sampling or data augmentation on the VAS scores classed as high by the average reader estimate we are likely to oversample from the least reliable part of the distribution with the most falsely labelled data-points and the lowest signal to noise ratio.

**Fig 14.**
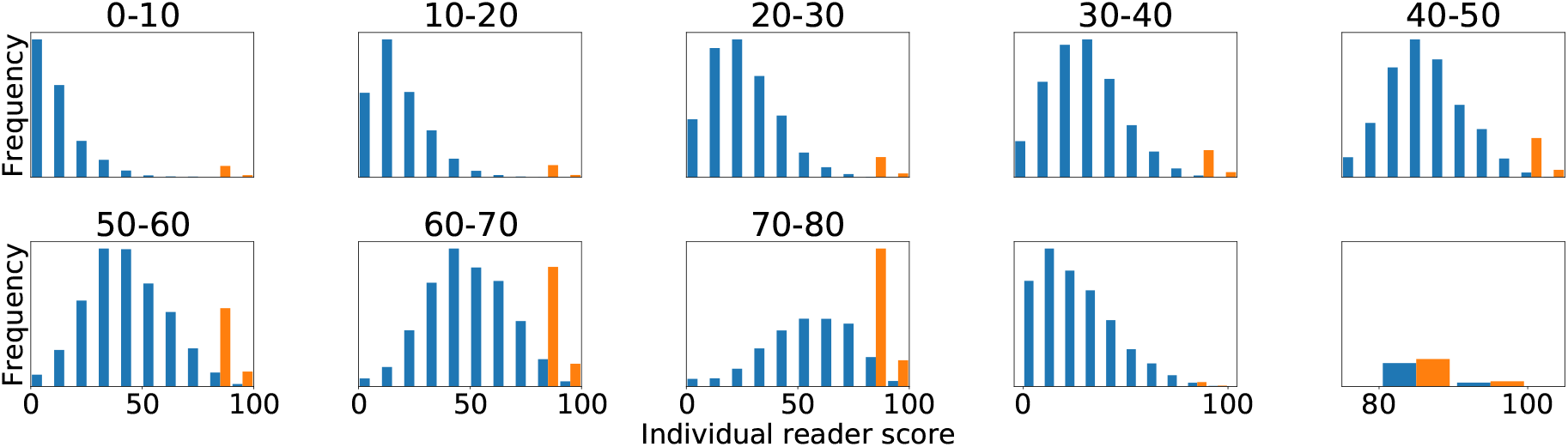
For each of the first eight deciles we show the distribution of the pair values as blue bars. For example, in the 1-10 decile range we show the distribution of the reader values which have a paired score between 1 and 10. We also show the number of reader averaged scores that are above 80. The bottom right two plots show the entire 0-80 distributions and a zoomed in version of the same results.

In Figure 15 we show plots of the pairs of VAS labels for the modelled and real estimates, we show 3,000 random pairs so that the structure of the distribution is visible. The dotted line shows a perfect relationship between the pair of readers. The left plot is the lower error (higher correlation) end of our modelled range and the right plot our higher error (lower correlation) modelled range. In the middle we show the real pairs of reader labels plotted next to one another, these points are very slightly perturbed so individual points can be seen.

**Fig 15.**
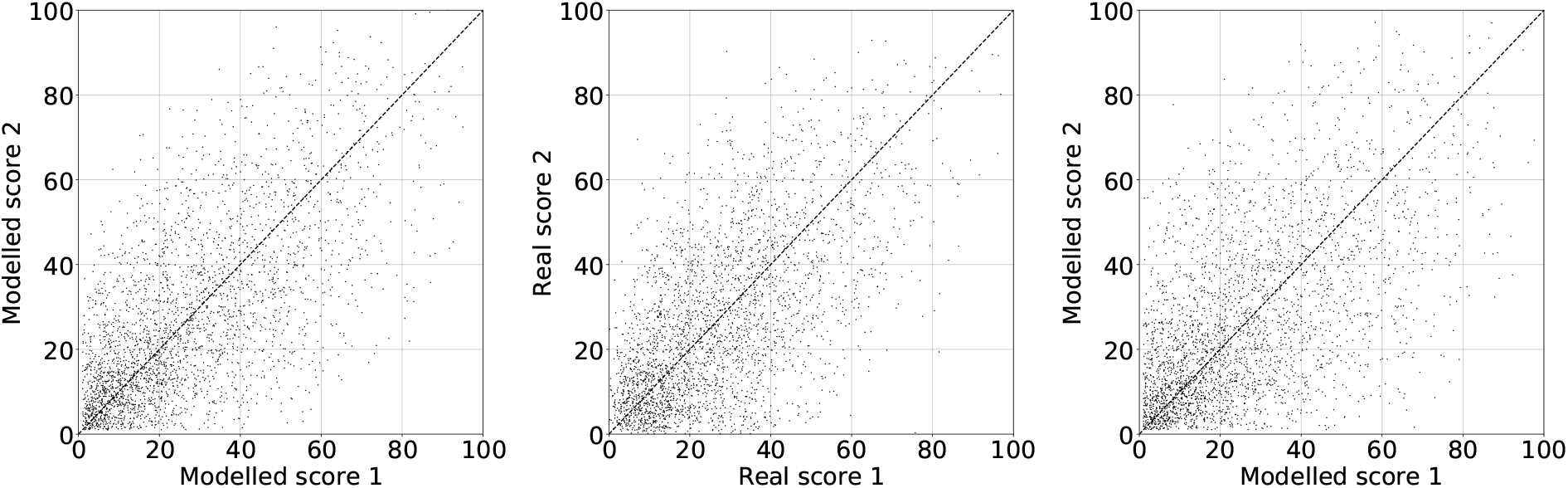
Examples of the pairs of scores for the modelled and real data, the dotted line shows a perfect relationship between the pair of estimates. Left) The modelled pairs of scores for the results produced with the lowest errors. Middle) The pairs of real reader scores. Right) The modelled pairs of scores produced with the highest error in the range.

In Figure 16 we show the average modelled optimal scores versus the average of the two modelled labels. The left plot is a direct comparison and the right plot a Bland-Altman plot for the difference between the modelled optimal score and the modelled average reader scores versus the average of the optimal and modelled scores.

**Fig 16.**
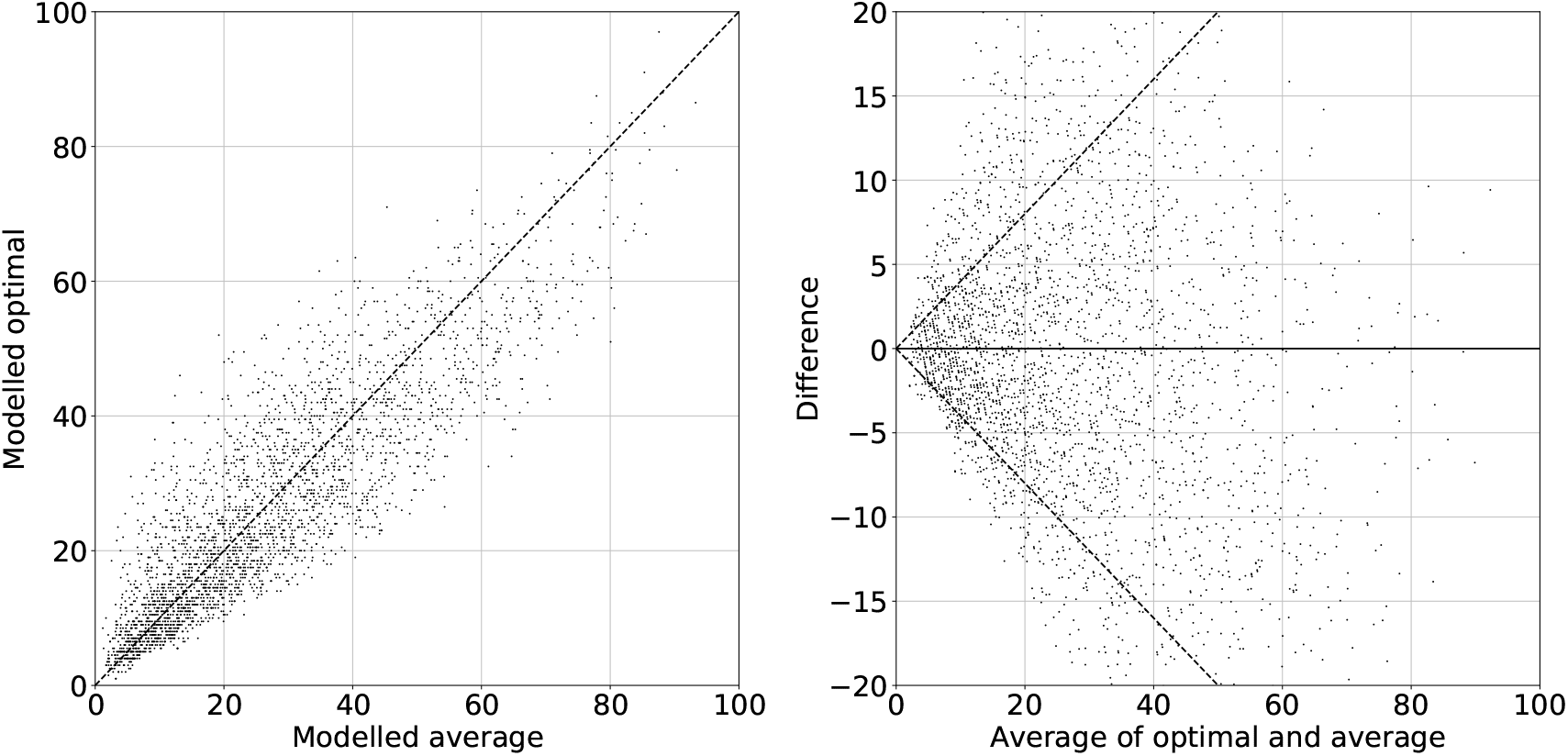
Modelled optimal scores and modelled reader average scores. Left) Direct relationship between the modelled optimal scores versus the averaged modelled pairs of estimates. Right) Bland-Altman plots of the difference between the modelled optimal and modelled reader averaged scores versus the average of the modelled optimal and modelled reader average scores.

Previous work has shown that VAS density scores show a strong correlation to the risk of developing cancer.^4^ In this paper, we have demonstrated that the variability of the reader estimates means that we have to be cautious about the level of confidence we have in results derived from these labels. We therefore repeat the analysis for the Prior data-set from that previous work to calculate the odds ratios.^4, 15^ In addition we provide an additional piece of analysis by perturbing the averaged scores. The purpose of doing so is to give some intuition about how much the odds ratios might change with small variations in the reader scores. We added a random amount to all the averaged scores by sampling from a Gaussian distribution with three different standard deviations of: *σ* = 1, 2, 5. We re-sampled any scores that went above 100 or below 1, until we got a score within the correct range. We performed each set of perturbations 5 times. The perturbation we apply to the reader averaged scores are small when compared with the variability between the pairs of reader scores, which have an RMSE between readers of 16.2.

The results of the range of the odds ratios found are shown in Figure 17. We show odds ratio (OR) plots for the second (*Q*2), third (*Q*3), fourth (*Q*4) and fifth (*Q*5) quintiles. On the left side of the plots, labelled with *Orig*., is the non-perturbed odds ratios. The next five, labelled with *σ* = 1 show the five repeats when the perturbation is parameterized with a standard deviation of 1. The results for *σ* = 2 and *σ* = 5 are shown in the next two sections, separated by the dashed line. The crosses show the odds ratios and the error bars the upper and lower 95% confidence intervals found via bootstrapping.

**Fig 17.**
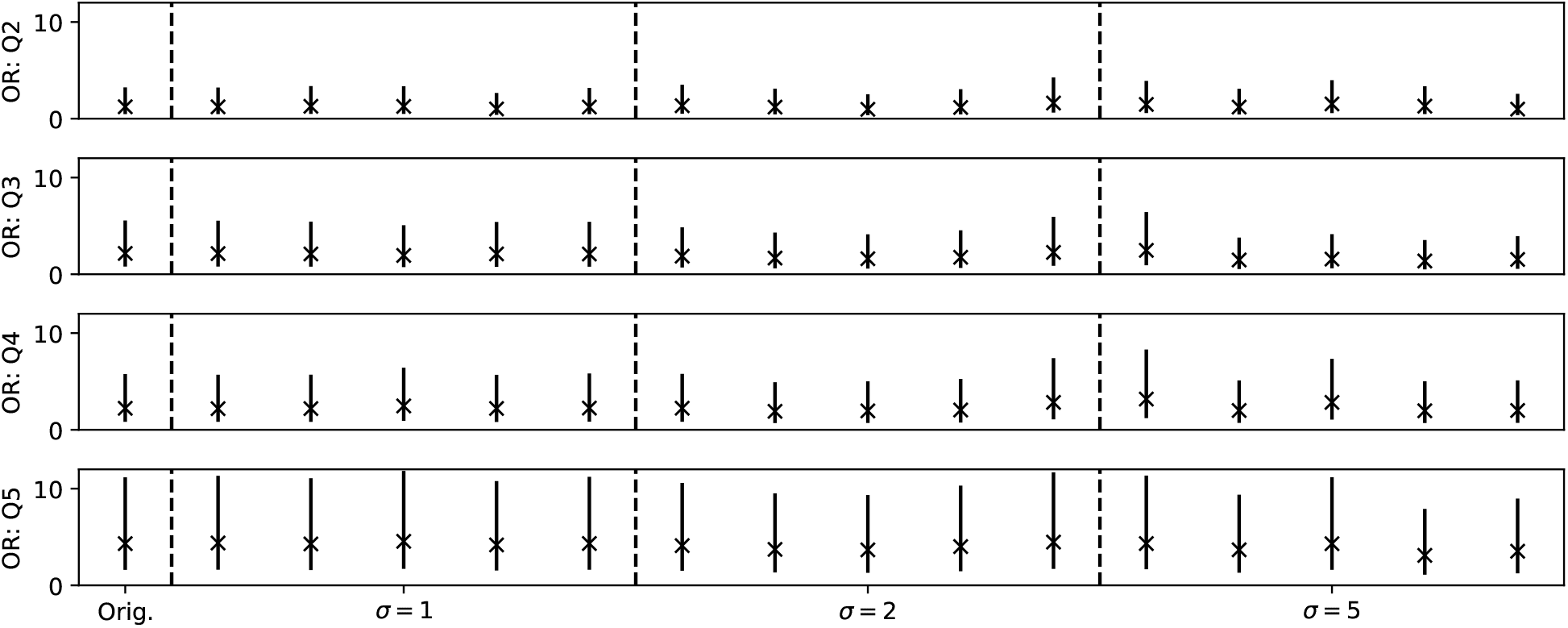
Plots of the odds ratios calculated using the reader averaged scores with uncertainties found via bootstrapping. *OR: Q2,Q3,Q4,Q5* shows the odds ratios for the second, third, fourth and fifth quintile respectively. *Orig*. shows the results with no perturbation, the five points around *σ* = 1, 2, 5 show the odds ratios when the reader averaged scores are perturbed by the standard deviation specified, with the multiple points being repeated applications of the perturbations across the data-set.

We therefore consider two aspects of uncertainty here: via bootstrapping we see the uncertainty relating to the sampling of the data and via the perturbations we see uncertainty due to the unreliability of the labels due to reader variability. The uncertainty due to sampling (the bootstrap error bars) are large and show the wide range of possible ORs that could occur with a different sample. The perturbations alter both the odds ratio and the uncertainty of it. We will see in the next section that the odds ratios found by the models are comparable with these odds ratios for the readers.

While this is a positive result for our models the problem is that the high level of uncertainty in the odds ratio values means we cannot easily assess whether our models are performing better than one another. This means, that we cannot bypass the metrics we considered earlier in this section to assess the quality of our models by looking directly at cancer risk, because the uncertainty involved is too large. From these perturbation results we would not want to trust that one model is better than another without quite significant differences in estimates. However, what this also shows is that the VAS scores do produce a robust set of cancer risk prediction. In previous work the other measures of density studied did not produce a ratio of above 3.^4^ Therefore, although we cannot be confident that results in a fairly broad range are an improvement on other results we can be reasonably confident in the overall ability of VAS to make good risk estimates in comparison to other density scores.

### B.2 Model Predictions

In Table 7 we show the prediction results for the MLO images equivalent to Table 3.

**Table 7.** Comparison metrics between our models and the average labelled data for the MLO images.

| Model | Label | Obj. func. | Corr | RMSE | AME | Median |
| --- | --- | --- | --- | --- | --- | --- |
| ResNet | Average | LinReg | $0.81 \pm 0.01$ | $9.49 \pm 0.22$ | $7.32 \pm 0.17$ | $5.87 \pm 0.18$ |
| | | L1 | $0.82 \pm 0.01$ | $9.24 \pm 0.22$ | $7.02 \pm 0.16$ | $5.54 \pm 0.19$ |
| | | L1Smooth | $0.82 \pm 0.01$ | $9.35 \pm 0.23$ | $7.14 \pm 0.16$ | $5.69 \pm 0.2$ |
| | | MSE | $0.83 \pm 0.01$ | $9.13 \pm 0.21$ | $7.04 \pm 0.15$ | $5.60 \pm 0.16$ |
| | Individual | LinReg | $0.81 \pm 0.01$ | $9.48 \pm 0.22$ | $7.31 \pm 0.16$ | $5.88 \pm 0.18$ |
| | | L1 | $0.82 \pm 0.01$ | $9.25 \pm 0.23$ | $6.94 \pm 0.17$ | $5.34 \pm 0.2$ |
| | | L1Smooth | $0.82 \pm 0.01$ | $9.25 \pm 0.23$ | $6.95 \pm 0.16$ | $5.38 \pm 0.17$ |
| | | MSE | $0.83 \pm 0.01$ | $9.02 \pm 0.22$ | $6.89 \pm 0.15$ | $5.45 \pm 0.19$ |
| DenseNet | Average | LinReg | $0.83 \pm 0.01$ | $9.03 \pm 0.22$ | $6.96 \pm 0.15$ | $5.66 \pm 0.17$ |
| | | L1 | $0.83 \pm 0.01$ | $8.94 \pm 0.22$ | $6.75 \pm 0.16$ | $5.21 \pm 0.16$ |
| | | L1Smooth | $0.83 \pm 0.01$ | $8.94 \pm 0.23$ | $6.75 \pm 0.16$ | $5.22 \pm 0.17$ |
| | | MSE | $0.84 \pm 0.01$ | $8.82 \pm 0.22$ | $6.78 \pm 0.15$ | $5.33 \pm 0.17$ |
| | Individual | LinReg | $0.83 \pm 0.01$ | $9.04 \pm 0.21$ | $6.97 \pm 0.15$ | $5.66 \pm 0.16$ |
| | | L1 | $0.83 \pm 0.01$ | $9.01 \pm 0.24$ | $6.75 \pm 0.17$ | $5.10 \pm 0.18$ |
| | | L1Smooth | $0.83 \pm 0.01$ | $9.03 \pm 0.23$ | $6.75 \pm 0.16$ | $5.12 \pm 0.15$ |
| | | MSE | $0.83 \pm 0.01$ | $8.92 \pm 0.21$ | $6.77 \pm 0.15$ | $5.22 \pm 0.15$ |
| Ensemble | | L1 | $0.84 \pm 0.01$ | $8.68 \pm 0.21$ | $6.59 \pm 0.15$ | $5.12 \pm 0.17$ |

Plots of the final ensemble predictions versus labels per woman are displayed in Figure 18. There are 2,682 women in the test set who have all labels and all predictions intact, 15 women are missing labels or images and are removed. We see the same general pattern as the per-image plots of Figure 6.

**Fig 18.**
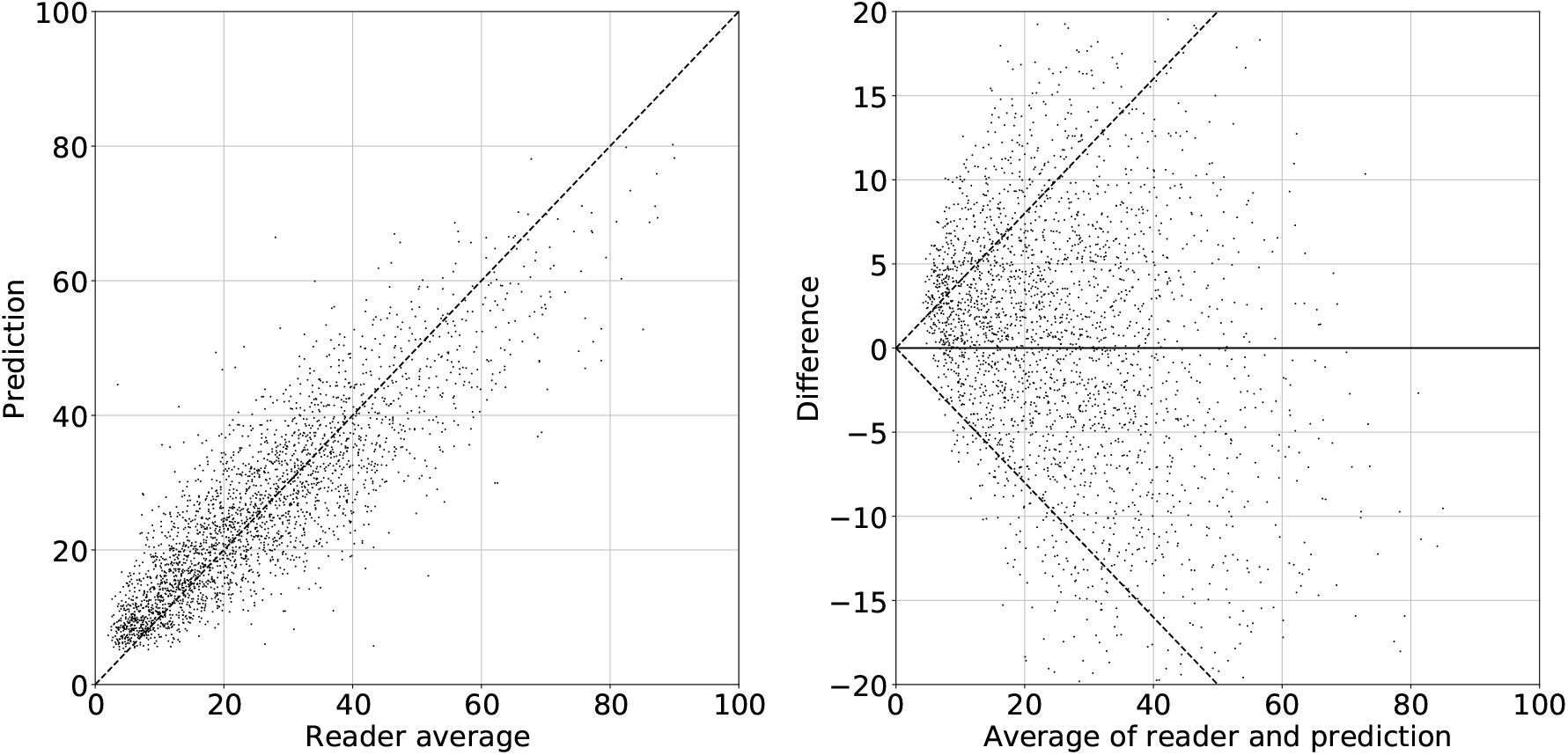
Density prediction per woman by the ensemble model compared to the averaged reader woman estimates. Left) Direct comparison between the ensemble model prediction and the reader average score. Right) Bland-Altman plots of the difference between the ensemble model prediction and the reader average score against the average of the two.

## Acknowledgements

D. Gareth Evans, Elaine Harkness and Susan M Astley are supported by the National Institute for Health Research (NIHR) Manchester Biomedical Research Centre (IS-BRC-1215-20007).

